# Explainable Artificial Intelligence for Prognostic Stratification in Out-of-Hospital Cardiac Arrest Patients Undergoing Extracorporeal Cardiopulmonary Resuscitation

**DOI:** 10.1101/2025.10.07.25337539

**Authors:** Yusuke Watanabe, Hirohiko Kohjitani, Yoshinori Matsuoka, Toshiaki Toyota, Madoka Sano, Yuta Azumi, Hideyuki Hayashi, Ryosuke Murai, Junichi Ooka, Yasuhiro Sasaki, Tomohiko Taniguchi, Kitae Kim, Atsushi Kobori, Natsuhiko Ehara, Makoto Kinoshita, Akihiko Inoue, Toru Hifumi, Tetsuya Sakamoto, Yasuhiro Kuroda, Yosuke Yamamoto, Koichi Ariyoshi, Yasushi Okuno, Yutaka Furukawa, Koh Ono, the SAVE-J II study group

**Author notes:** Corresponding author contact information: Hirohiko Kohjitani, Department of Cardiovascular Medicine, Graduate School of Medicine, Kyoto University, 54 Shogoin Kawahara-cho, Sakyo-ku, Kyoto, Kyoto, 606-8507, Japan., Toshiaki Toyota, Department of Cardiovascular Medicine, Kobe City Medical Center General Hospital 2-1-1, Minatojima-minamimachi, Chuo-ku, Kobe, Hyogo, 650-0047, Japan.

## Abstract

**Background and Aims:** Prognostication in patient with out-of-hospital cardiac arrest (OHCA) underwent extracorporeal cardiopulmonary resuscitation (ECPR) remains challenging due to the complexity of clinical variables. We aimed to develop and interpret artificial intelligence (AI) models for early outcome prediction in OHCA patients treated with ECPR, and to identify clinically meaningful patient subgroups through supervised clustering based on model explanations.

**Methods:** We retrospectively analyzed data from the SAVE-J II registry, a multicenter registry of adult OHCA patients treated with ECPR in Japan. We defined and developed prediction models for all-cause death: Cohort 1 included all patients for predicting day 1 outcomes using binary classification models, and Cohort 2 excluded patients who died on day 1 deaths and developed survival models for events from day 2 onward. Models were interpreted using Shapley Additive exPlanations (SHAP), and hierarchical clustering based on SHAP values was performed to stratify patients into prognostic subgroups.

**Results:** In cohort 1 (n=1,624, age 60 IQR [49-68]), 433 (26.7%) all-cause death occurred on day 1, and AI models achieved 0.85 of AUC. In cohort 2 (n=1,191, age 59 IQR [48-67]), 752 (63.1%) all-cause deaths occurred from day 2. AI models achieved a mean of time-dependent AUCs of 0.77. SHAP analysis identified different predictive variables between cohorts. SHAP-based hierarchical clustering revealed patient groups with markedly different prognoses.

**Conclusions:** AI models accurately predicted short-term outcomes in ECPR-treated OHCA patients and revealed temporal shifts in key prognostic factors. SHAP-based clustering enabled meaningful stratification and may support personalized treatment strategies.

**Structured graphical abstract:** *Key Question:* Can AI models accurately predict all-cause death in patients who underwent ECPR (Extracorporeal cardiopulmonary resuscitation) for OHCA (out-of-hospital cardiac arrest) and can SHAP (Shapley Additive Explanations) values reveal clinically meaningful patient subgroups?

*Key Finding:* AI models accurately predicted all-cause mortality, though less so for bleeding. Landmarking patients at day 1 and interpreting the models with SHAP values revealed differing early and later event characteristics. SHAP-based supervised clustering stratified patients into prognostically distinct groups.

*Take-home Message:* By employing interpretable AI models, patient prognoses can be estimated while elucidating the underlying factors. AI models will help clinicians make treatment decisions for patients who underwent ECPR. 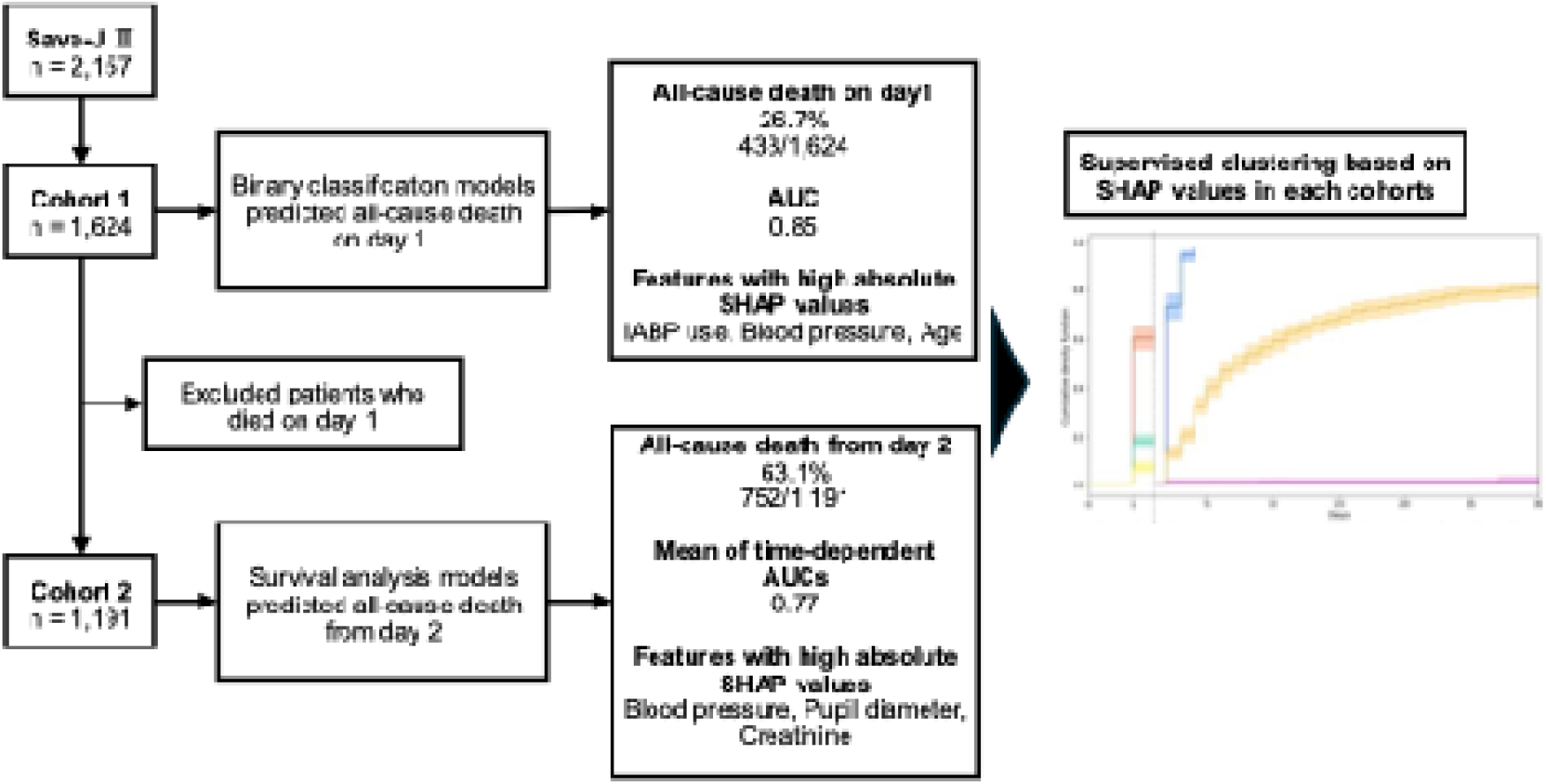

## Introduction

Out-of-hospital cardiac arrest (OHCA) is a significant public health concern, with high incidence and mortality rates worldwide. Despite advancements in resuscitation strategies, overall survival to hospital discharge remains poor, especially in patients with refractory cardiac arrest^1^. Extracorporeal cardiopulmonary resuscitation (ECPR), which combines conventional cardiopulmonary resuscitation with extracorporeal membrane oxygenation (ECMO), has emerged as a promising salvage therapy for selected patients with refractory OHCA^2–4^. Although recent studies have suggested potential survival and neurological benefits, outcomes remain highly variable. This variability arises from heterogeneity in patient characteristics, arrest circumstances, and institutional practices, highlighting the need for improved prognostic tools to support individualized treatment decisions in ECPR.

Accurate early prognostication is particularly challenging in this setting^5–7^. Traditional prognostic scoring systems, such as the TiPS65 score^6^, offer simple and rapid bedside assessment but are limited by their reliance on a small number of variables and their inability to capture complex interactions inherent in critical care. Machine learning (ML) models have demonstrated superior predictive performance by leveraging high-dimensional clinical data and modeling non-linear relationships^8,9^. However, their clinical utility has been constrained by poor interpretability—so-called “black-box” behavior.

To bridge this gap, explainable artificial intelligence (XAI) techniques, notably Shapley Additive Explanations (SHAP), have been developed to attribute individual model predictions to contributing variables^10^. This interpretability can enhance clinicians’ trust in model outputs and facilitate integration into bedside decision-making. Nevertheless, the application of XAI in ECPR remains limited, and few studies have employed these tools in large, real-world cohorts.

Prior efforts to predict outcomes in patients undergoing ECPR using ML have shown promise but remain methodologically limited. For instance, the ECMO Predictive Algorithm (ECMO PAL), trained on the international ELSO registry, demonstrated improved in-hospital mortality prediction in venoarterial ECMO patients^11^. However, its applicability to OHCA-ECPR patients is unclear due to a lack of specific validation. Single-center studies, such as those by Crespo-Diaz et al^9^, have achieved moderate discrimination in OHCA-ECPR cohorts but lack external generalizability. Moreover, most prior models lack explainability, limiting their clinical implementation.

To address these limitations, we aimed to develop and interpret explainable ML models to predict early mortality and bleeding complications in OHCA patients undergoing ECPR. Leveraging the SAVE-J II registry—a large, multicenter cohort in Japan—we constructed predictive models using algorithms such as Light Gradient Boosting Machine (LightGBM) and Gradient Boosting Survival Analysis (GBSA). To ensure transparency, we applied SHAP to interpret individual predictions and identify key prognostic features. Furthermore, we performed hierarchical clustering on SHAP values to uncover clinically meaningful subgroups. This approach aims to provide a robust, interpretable, and generalizable framework for outcome prediction and risk stratification in ECPR-treated OHCA patients.

## Methods

### Study design and participants

The SAVE-J II registry is a multicenter retrospective study conducted in Japan at 36 participating institutions^12^. Between January 2013 and December 2018, this study enrolled OHCA patients aged 18 years or older who received ECPR at the respective emergency department. The exclusion criteria for the current analysis were as follows: patients who received VA-ECMO after ICU admission; patients who withdrew after cannulation due to return of spontaneous circulation (ROSC); patients with external causes of cardiac arrest such as acute aortic dissection/aortic aneurysm, hypothermia, primary cerebral disorders, infection, drug intoxication, trauma, suffocation, and drowning; patients who achieved ROSC upon hospital arrival and ECMO initiation; patients transferred from other hospitals; patients with unknown outcomes and cannula insertion failure; and patients without records of bleeding complications.

To examine temporal differences in clinical outcomes and decision-making after ECPR initiation, we created two cohorts from the SAVE-J II registry. After applying the exclusion criteria, the patients remaining in the registry were included in Cohort 1. From this group, we excluded patients who died on day 1, thus forming Cohort 2.

The design and data collection methods of the SAVE-J II study have been described previously^12^. Data collected included patient demographics, prehospital events, clinical interventions, and outcomes. Performance status (PS) before admission (a lower PS score indicates a higher level of performance of daily activities: a score of 0 indicates that the patient is fully active and able to carry out all activities without restriction, while a score of 4 indicates that the patient is completely disabled and bedridden). Echo-guided puncture is defined as vessel puncture performed under real-time ultrasound visualization. The arrival-ECMO pump-on time was defined as the duration from the arrival of emergency medical services to the time of ECMO initiation, marked by the ECMO pump-on moment. Cardiac arrest was attributed either to cardiac causes, such as acute coronary syndrome, arrhythmias, cardiomyopathy, and myocarditis, or to non-cardiac causes, such as pulmonary embolism. Bleeding events that occurred during hospitalization were also assessed, defined as those requiring blood transfusion, interventional radiology (IVR), or surgical intervention for hemostasis, excluding cerebral hemorrhage verified by CT. In-hospital death was defined as mortality from any cause during hospitalization.

### Statistical analysis

Categorical variables are presented as numbers and percentages. Continuous variables are expressed as medians with interquartile ranges (IQR). The participating institutions were classified into four groups (Q1 to Q4) based on the quartiles of ECPR case volumes in descending order. The cumulative incidence of all-cause death was estimated using the Kaplan–Meier method to assess the differences among clusters within each cohort.

### Development of prediction models

Prediction models for all-cause deaths and bleeding complications were developed using information obtained on the first day of admission. We used only explanatory variables with missing values below 30% to develop the models. All explanatory variables were imputed for missing values before use in the model development. Missing values were imputed using multivariate feature imputation, which is an imputation method that uses all other variables as well as those with missing values^13^. In the prediction models for Cohort 2, bleeding, infarction, and infection events that occurred on day 1 were added as explanatory variables.

In Cohort 1, binary classification models were developed for all-cause deaths and bleeding events on day 1. A Light Gradient-Boosting machine (LightGBM), random forest (RF), and extreme gradient-boosting (XGBoost) were applied to Cohort 1^14–16^. In Cohort 2, a survival analysis model was constructed for all-cause death and bleeding events from day 2 onward. Gradient-boosting survival analysis (GBSA), random survival forest (RSF), and extra survival trees (EST) were utilized as survival models in Cohort 2^17–19^.

In both cohorts, the original dataset was randomly split into training and test datasets in a 4:1 ratio to maintain consistent rates of all-cause death and bleeding events between the two groups. The hyperparameters of all the models were tuned using Bayesian optimization with stratified five-fold cross-validation, which is a widely recognized automated technique for hyperparameter tuning in AI models^20^. All the prediction models were developed based on the training dataset, and their performances were evaluated using the test dataset.

### Evaluation of models and XAI application with SHAP

The performance of the models in both cohorts was evaluated using a test dataset. In Cohort 1, the model performance was evaluated based on the area under the receiver operating characteristic curve (AUC). The optimal cutoff value was determined by identifying the maximum Youden index, which was subsequently used to compute sensitivity and specificity. In Cohort 2, the models were evaluated using the mean time-dependent AUC, which was defined as the average AUC calculated daily from days 2 to 30.

To understand the reasoning behind the predictions made by the complex AI models, additional analyses were performed using SHAP, a unified framework for interpreting machine learning (ML) predictions^19^. The absolute SHAP values of the relevant variables indicate the magnitude of their influence on the prediction outcome. The SHAP values can be either positive or negative. A higher positive SHAP value indicates a greater positive impact on the prediction outcome, whereas a higher negative SHAP value indicates a greater negative impact. In this study, the SHAP values were calculated to visualize the impact of each variable on each model.

In the SHAP summary plots, the feature ranking (y-axis) indicates the variables that are important for the prediction outcome and the SHAP value (x-axis) is a unifying indicator of the influence of each variable on the model. Each variable is indicated by differently colored dots for the attribution of all patients to the predicted outcome. For the numerical variables, low actual values are represented by blue dots, whereas high actual values are represented by red dots. For categorical variables processed with one-hot encoding, blue and red dots represent 0 and 1, respectively.

Using hierarchical clustering based on the SHAP values, the patients were classified into clusters. A dendrogram was created using the Euclidean method and appropriate cluster divisions were created. For each cluster, the top 20 features based on mean absolute SHAP values were compared. Cumulative incidence curves for all-cause deaths were generated to assess the prognoses of each cluster.

### Hierarchical clustering based on SHAP values

Hierarchical clustering was performed based on the SHAP values of the 20 variables with the highest mean absolute SHAP values. Based on the clustering results, we classified appropriate clusters and evaluated the prognoses of each cluster by calculating the cumulative event incidence. For comparison with clustering based on the SHAP values, we standardized the actual measurements of the same 20 variables and conducted hierarchical clustering based on an identical method.

### Ethical statement

The current analysis was performed as a part of the data from SAVE-J II registry, which was registered at the University Hospital Medical Information Network Clinical Trials Registry and the Japanese Clinical Trial Registry (registration number: UMIN000036490), approved by the institutional review board of Kagawa University (approval number: 2018–110) and each participating institution, including Kobe City Medical Center General Hospital (approval number: zn200304). Informed consent from the patient was waived owing to the retrospective nature of the study design, and all procedures were performed in accordance with the ethical standards of the review board of Kobe City Medical Center General Hospital on human experimentation and with the Declaration of Helsinki of 1975.

### Package for analysis

The ML model was developed using the following libraries and tools: Python (3.9.19), NumPy (1.26.4), Pandas (2.2.2), Scikit-learn (1.4.2), XGBoost (2.0.3), LightGBM (4.3.0), Matplotlib (3.9.0), Seaborn (0.13.2), SHAP (0.45.1), Optuna (3.6.1), Scikit-survival (0.23.0), Scikit-optimize (0.10.2), and Lifelines (0.29.0).

### Model Availability

Main machine lerning models and initial codes are available online at https://github.com/clinfo/ML_SAVE_J2

## Results

### Patient characteristics

Figure 1 shows a flowchart depicting the eligibility of the patients for analysis. During the study period, data from 2,157 patients were included in the SAVE-J II registry. A total of 535 patients were excluded. Thereafter, the remaining patients were assigned to Cohort 1, consisting of 1,624 patients. Cohort 2 was further refined by excluding 433 patients who died on days 1, leaving 1,191 patients.

**Figure 1.**
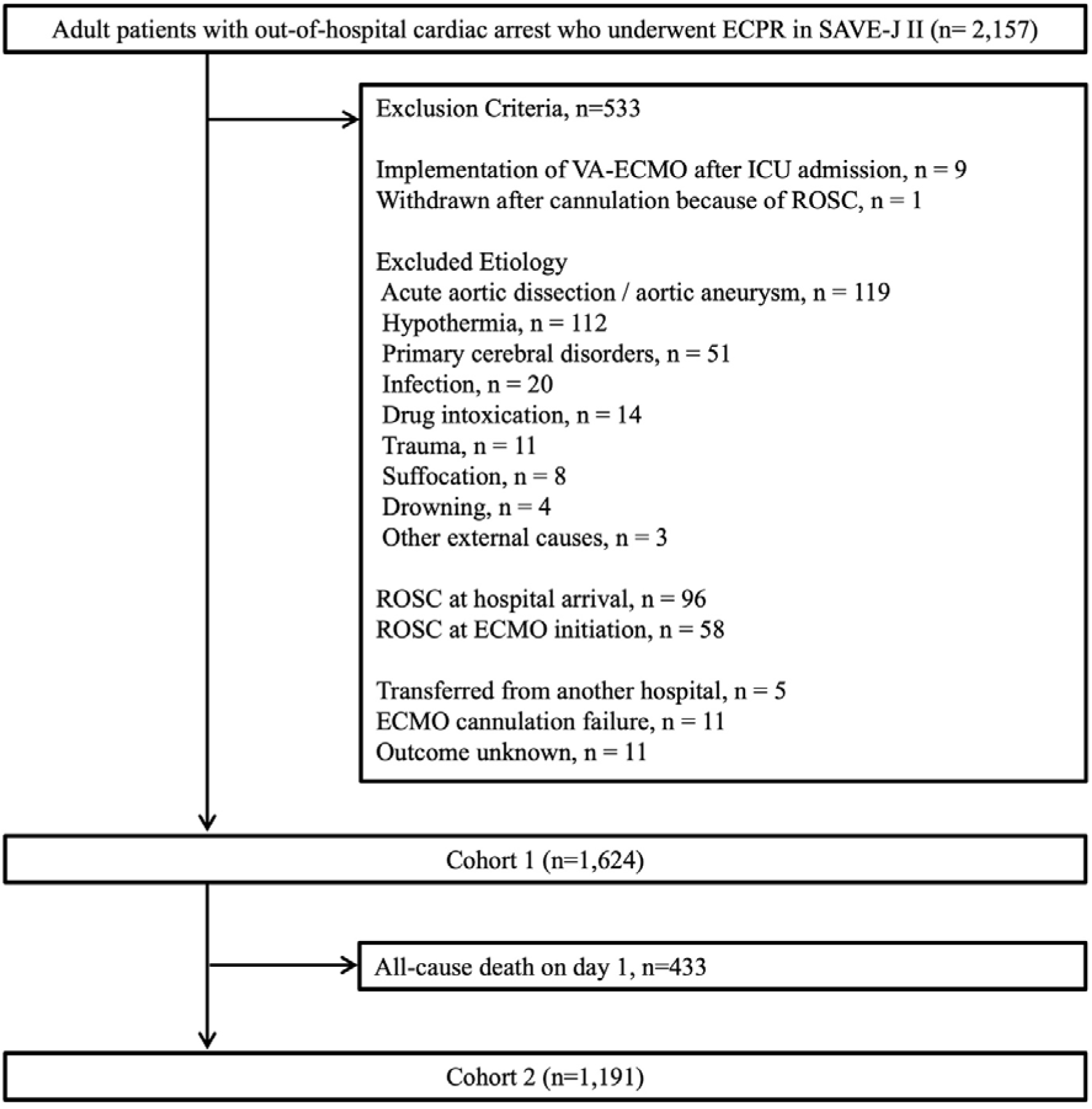
Study flow chart. ECPR, extracorporeal cardiopulmonary resuscitation; VA-ECMO, venoarterial extracorporeal membrane oxygenation; ICU, intensive care unit; ROSC, return of spontaneous circulation

Cohort 1 included patients with a median age of 60 [IQR: 49–68] years, of whom 84.6% were male. In this cohort, all-cause deaths occurred in 433 patients (26.7%), while bleeding on the first day was observed in 206 patients (12.7%). Cohort 2 consisted of 1,191 patients, with 83.7% being male and a median age of 59 [IQR: 48–67] years. In this cohort, the median observation period for all-cause deaths was 7 days [IQR, 2–28], during which 752 patients died (63.1%). Bleeding beyond day 2 was observed in 108 patients (9.1%) over a median duration of 5 days [IQR: 1–24 days].

### Model performance

#### Cohort 1

In Cohort 1, among the three AI models (LightGBM, RF, and XGBoost) evaluated, LightGBM demonstrated the best performance in predicting all-cause deaths on day 1, with an AUC of 0.85, followed by RF (AUC = 0.83) and XGBoost (AUC = 0.79). Using the LightGBM model in the test set (Fig. 1a) with an optimal cutoff of 0.36 determined by the Youden index, the model achieved an accuracy of 0.82, sensitivity of 0.71, specificity of 0.86, precision of 0.65, and negative predictive value of 0.89. For day 1 bleeding prediction, the LightGBM model showed the highest performance (AUC = 0.56), outperforming RF (AUC = 0.55) and XGBoost (AUC = 0.47). In the test set (Fig. 1b), the LightGBM model with an optimal threshold of 0.12 achieved an accuracy of 0.50, sensitivity of 0.71, specificity of 0.47, precision of 0.16, and negative predictive value of 0.92.

#### Cohort 2

In Cohort 2, we developed three survival models (GBSA, RSF, and EST) to predict all-cause deaths and bleeding events between days 2 and 30. For all-cause deaths prediction, the GBSA achieved the highest mean time-dependent AUC, defined as the mean of time-dependent AUC from day 2 to day 30, of 0.77 in the test set (Fig. 2c), followed by the RSF (mean time-dependent AUC = 0.75) and EST (mean time-dependent AUC = 0.75). Similarly, for bleeding prediction, the GBSA demonstrated superior performance, with a mean time-dependent AUC of 0.67 (Fig. 2d), whereas the RSF (mean time-dependent AUC = 0.56) and extreme survival trees (mean time-dependent AUC = 0.56) showed lower performance.

**Figure 2:**
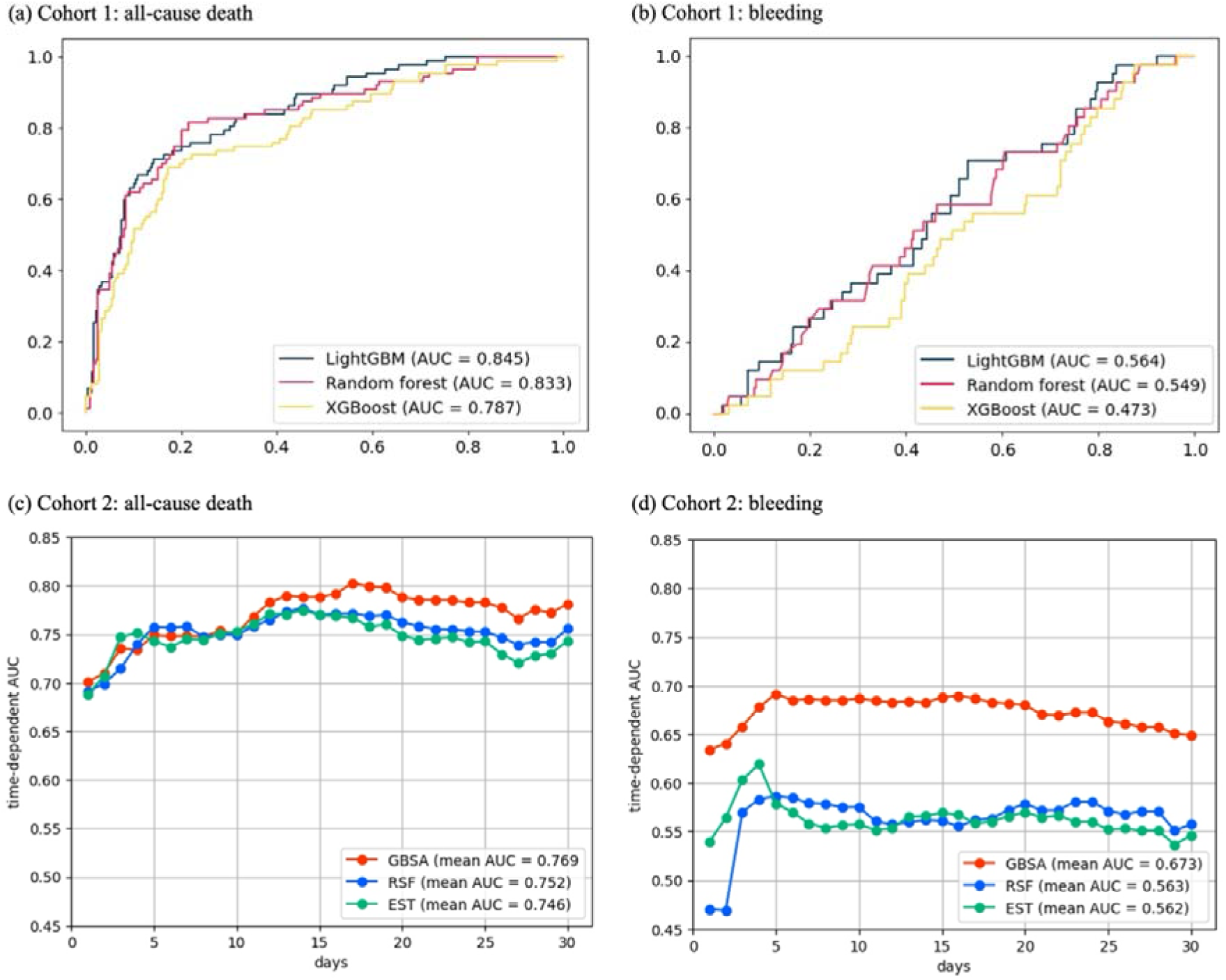
Performance of the models. Receiver operating characteristic (ROC) curve and time-dependent AUC for each model. (a) Receiver operating characteristic curve to predict bleeding on day 1 in Cohort 1 using LightGBM. (b) Time-dependent AUC to predict bleeding from day 2 after differentiating Cohort 2. (c) Receiver operating characteristic curve for predicting all-cause death on day 1 in Cohort 1 using LightGBM. (d) Time-dependent AUC to predict all-cause death from day 2 in Cohort 2. The optimal threshold was determined when the Youden index reached its maximum value. The time-dependent AUC represents the average value calculated from days 2 to 30. AUC, area under the ROC curve; EST, Extra Survival Trees; GBSA, Gradient-Boosting Survival Analysis; RSF, Random Survival Forest.

### Model interpretation through SHAP

Figure 2 illustrates the SHAP-based interpretation of the best-performing models to predict all-cause deaths and bleeding outcomes in Cohorts 1 and 2. In Cohort 1, feature plots from the best-performing model (LightGBM) identified important variables for all-cause deaths (Fig. 2a). The most predictive variables based on SHAP values for all-cause deaths were intra-aortic balloon pump (IABP) use, maintained blood pressure, age, potassium level and time from emergency medical service (EMS) arrival to ECMO pump-on (Fig. 2a). Many variables had a clear monotonic relationship with the outcomes. A lower likelihood of IABP use and advanced age were both consistently associated with increased mortality, indicating a negative impact on survival. In contrast, maintained blood pressure and a shorter duration from EMS arrival to ECMO pump-on were both consistently associated with improved survival (Fig. 2a).

In the GBSA of Cohort 2, the feature plots showed important variables in the all-cause deaths prediction model (Fig. 2c). The most predictive variables based on SHAP values were maintained blood pressure, pupil diameter, creatinine level, time from EMS arrival to ECMO pump-on, potassium, and white blood cell (WBC) count. A larger pupil diameter, longer time from EMS arrival to ECMO pump-on, and lower WBC count were monotonically associated with increased mortality from day 2 onwards. Conversely, higher creatinine levels gradually contributed to the mortality risk. The maintained blood pressure showed a clear monotonic relationship with mortality, whereas other variables demonstrated gradually evolving relationships. Notably, IABP use, which had the highest mean absolute SHAP value in Cohort 1, was not ranked among the top 20 variables in Cohort 2 (Fig. 2c).

Although the predictive accuracy for bleeding was limited, we calculated the SHAP values, as shown in Figures 2b and 2d. In Cohort 1, percutaneous coronary intervention (PCI) or IABP use was associated with an increased risk of bleeding, whereas in Cohort 2, the insertion of ECMO in the catheterization laboratory or patients with multivessel disease was associated with a higher risk of bleeding.

### Hierarchical clustering based on SHAP values

Hierarchical clustering based on the SHAP values for mortality prediction models was performed for both Cohort 1 and 2, it identified three clusters for each cohort (Fig. 4). Based on the mortality rates from highest to lowest, the clusters were labeled as clusters A, B, and C in Cohort 1 and clusters X, Y, and Z in Cohort 2. Analysis of the SHAP value patterns (Fig. 5) revealed that the clusters in Cohort 1 were primarily differentiated by IABP use and blood pressure, whereas the clusters in Cohort 2 were characterized by multiple correlated clinical variables. These distinctions were further confirmed by the cluster characteristics summarized in Table 2, where Cohort 1 clusters showed marked differences in IABP use and blood pressure (Table 2a), and Cohort 2 clusters demonstrated equally distributed changes across multiple clinical variables (Table 2b). The cumulative incidence of mortality for each cluster (Fig. 6) demonstrated that hierarchical clustering based on SHAP values identified clusters with markedly different prognoses compared to those obtained from unsupervised clustering (Supplemental Fig. 2).

**Figure 3.**
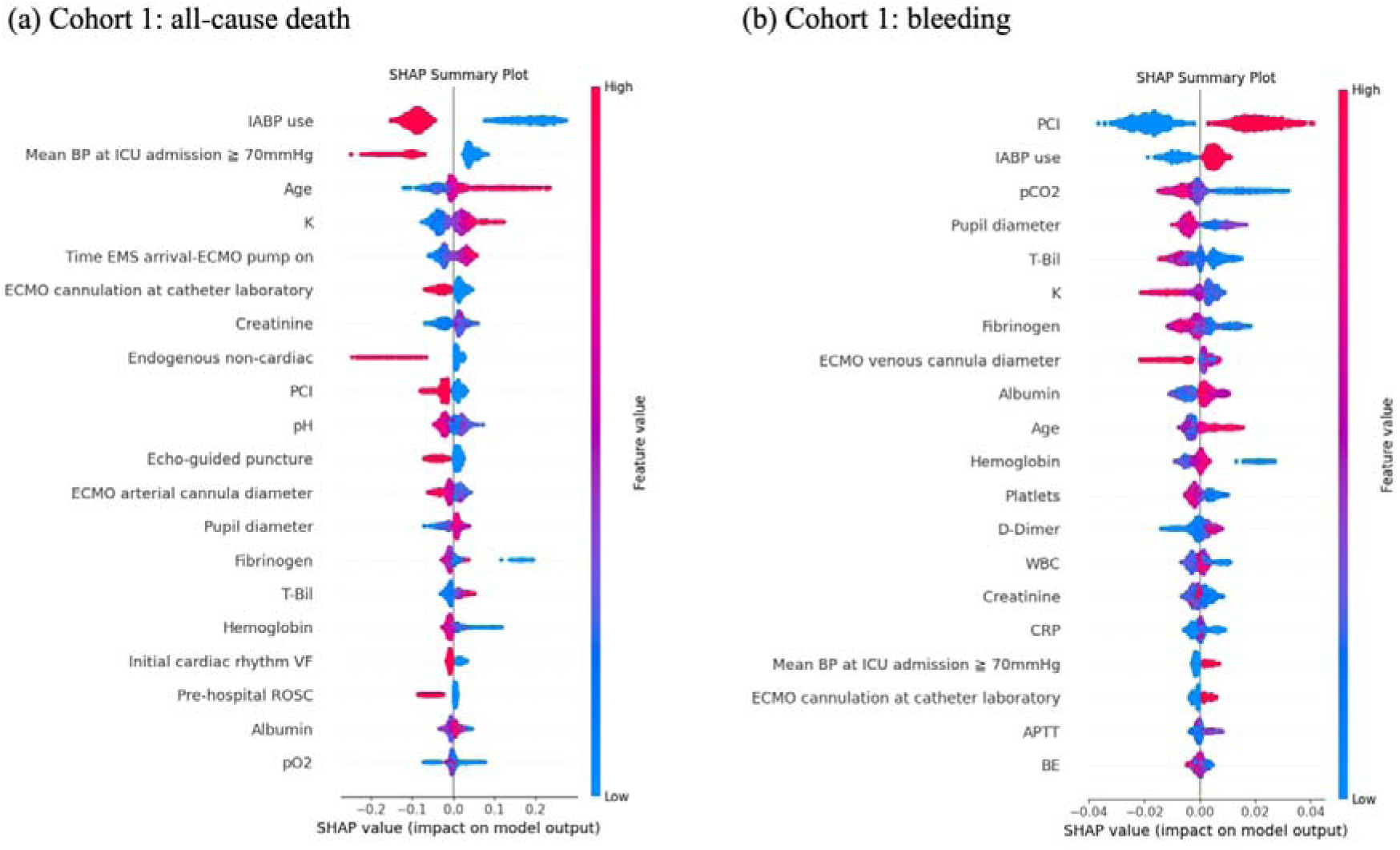
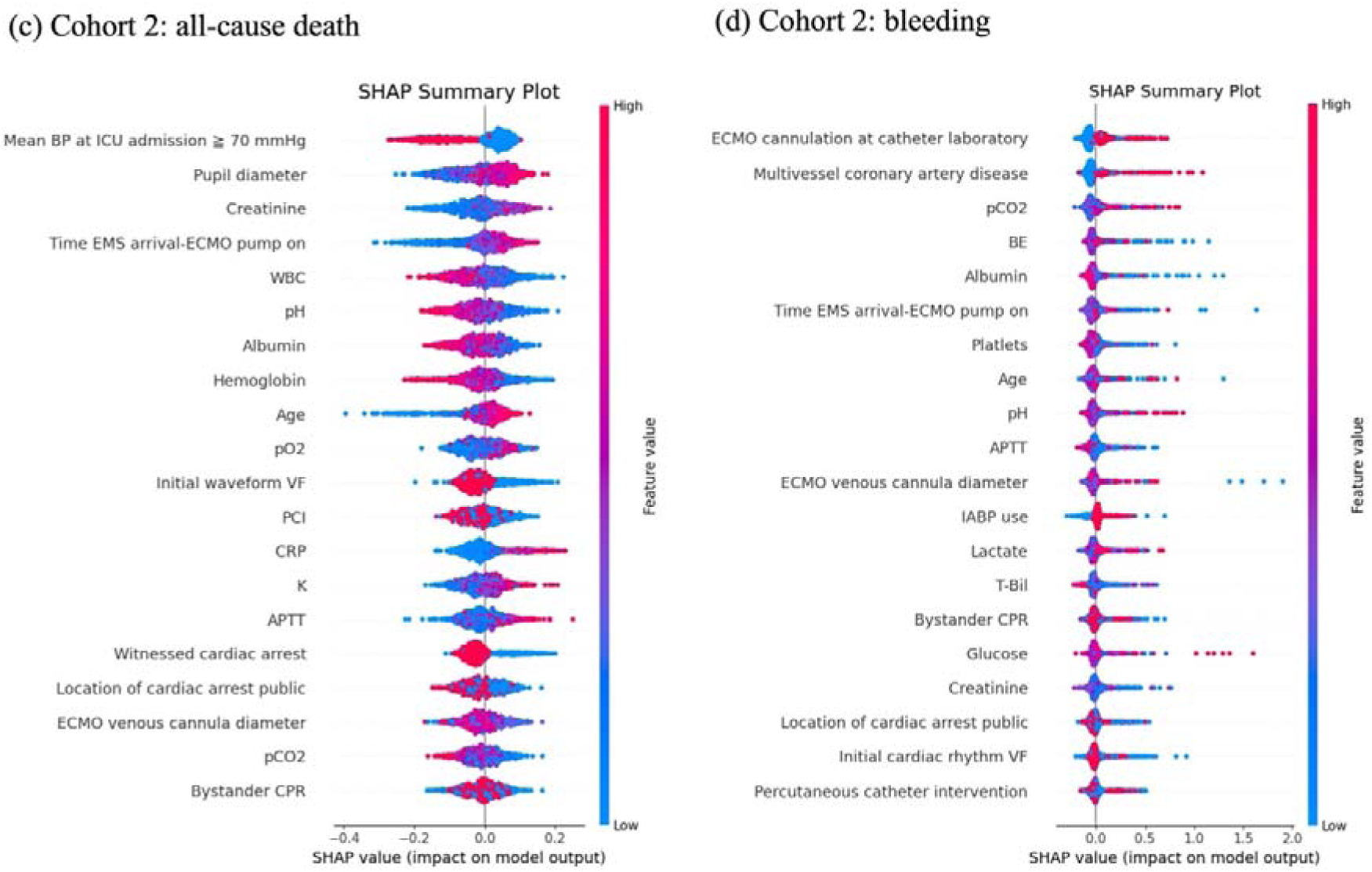
SHAP summary plot. Features indicating higher overall SHAP. A higher SHAP value for a given feature indicates a greater risk of death or bleeding. The red shading in the feature value signifies a higher value. (a) SHAP summary plot to predict bleeding on day 1 in Cohort 1 using the LightGBM model. (b) SHAP summary plot to predict all-cause death on day 1 in Cohort 1 using the LightGBM model. (c) The SHAP summary plot to predict bleeding from day 2 in Cohort 2 using the Gradient-Boosting Survival Analysis model. (d) SHAP summary plot to predict all-cause death from day 2 in Cohort 2 using the Gradient-Boosting Survival Analysis model. APTT, activated partial thromboplastin time; AUC, area under the ROC curve; BE, base excess; CRP, C-reactive protein; ECMO, extracorporeal membrane oxygenation; EMS, emergency medical services; IABP, intra-aortic balloon pump; ICU, intensive care unit; LightGBM, light gradient-boosting machine; PCI, percutaneous coronary intervention; ROSC, return of spontaneous circulation; T-Bil, total bilirubin; VF, ventricular fibrillation; WBC, white blood cell count.

**Figure 4:**
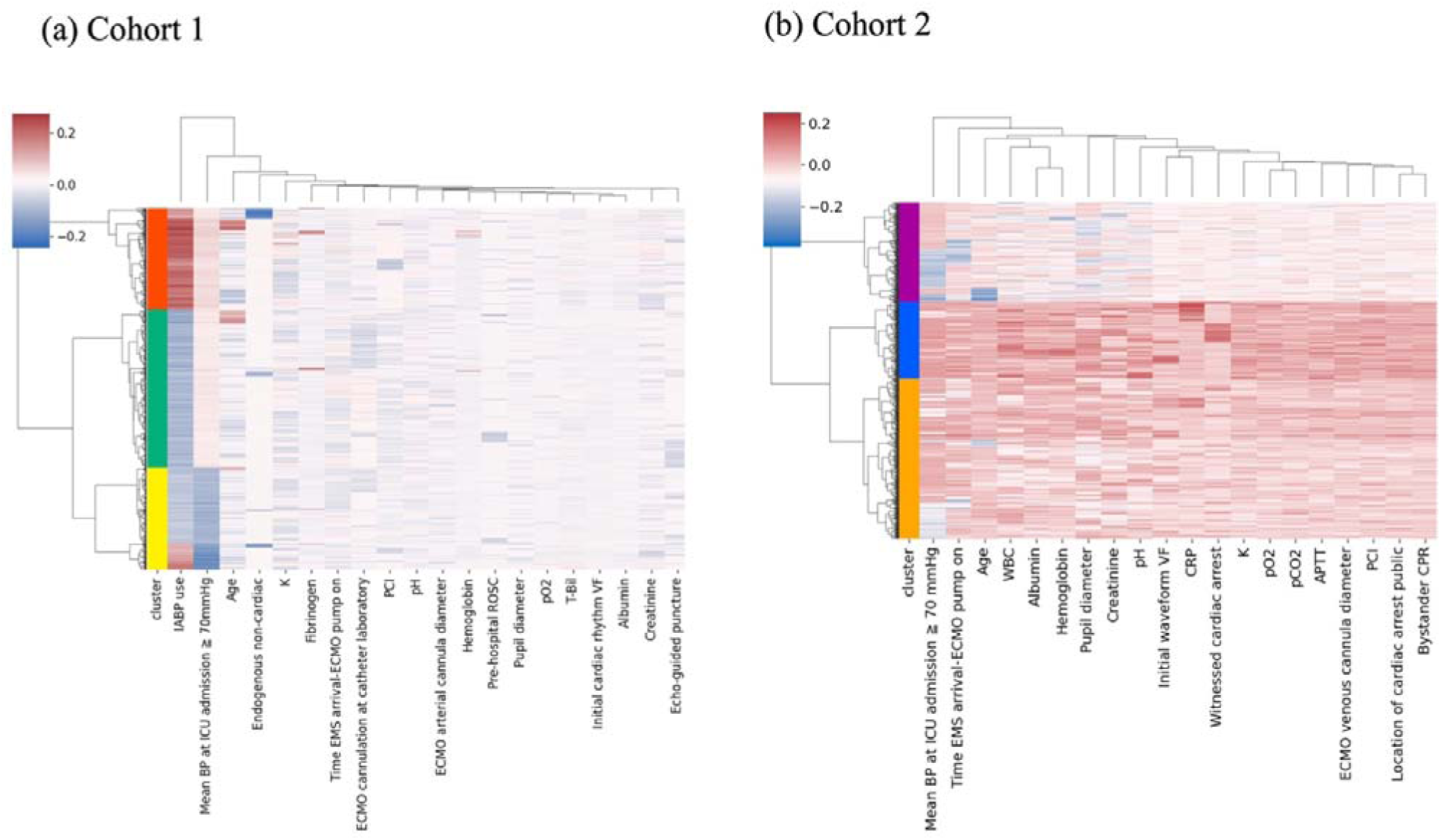
Heatmaps of hierarchical clustering based on SHAP values. Heatmaps displaying the results of hierarchical clustering based on SHAP values for Cohorts 1 and 2, to predict all-cause death. (a) Cluster map of SHAP values from the model predicting day 1 mortality in Cohort 1 developed using LightGBM. (b) Cluster map of SHAP values from the model predicting mortality from day 2 in Cohort 2 developed using gradient-boosting survival analysis. APTT, activated partial thromboplastin time; AUC, area under the ROC curve; CRP, C-reactive protein; ECMO, extracorporeal membrane oxygenation; EMS, emergency medical services; IABP, intra-aortic balloon pump; ICU, intensive care unit; LightGBM, light gradient-boosting machine; PCI, percutaneous coronary intervention; ROSC, return of spontaneous circulation; VF, ventricular fibrillation.

**Table 1:**
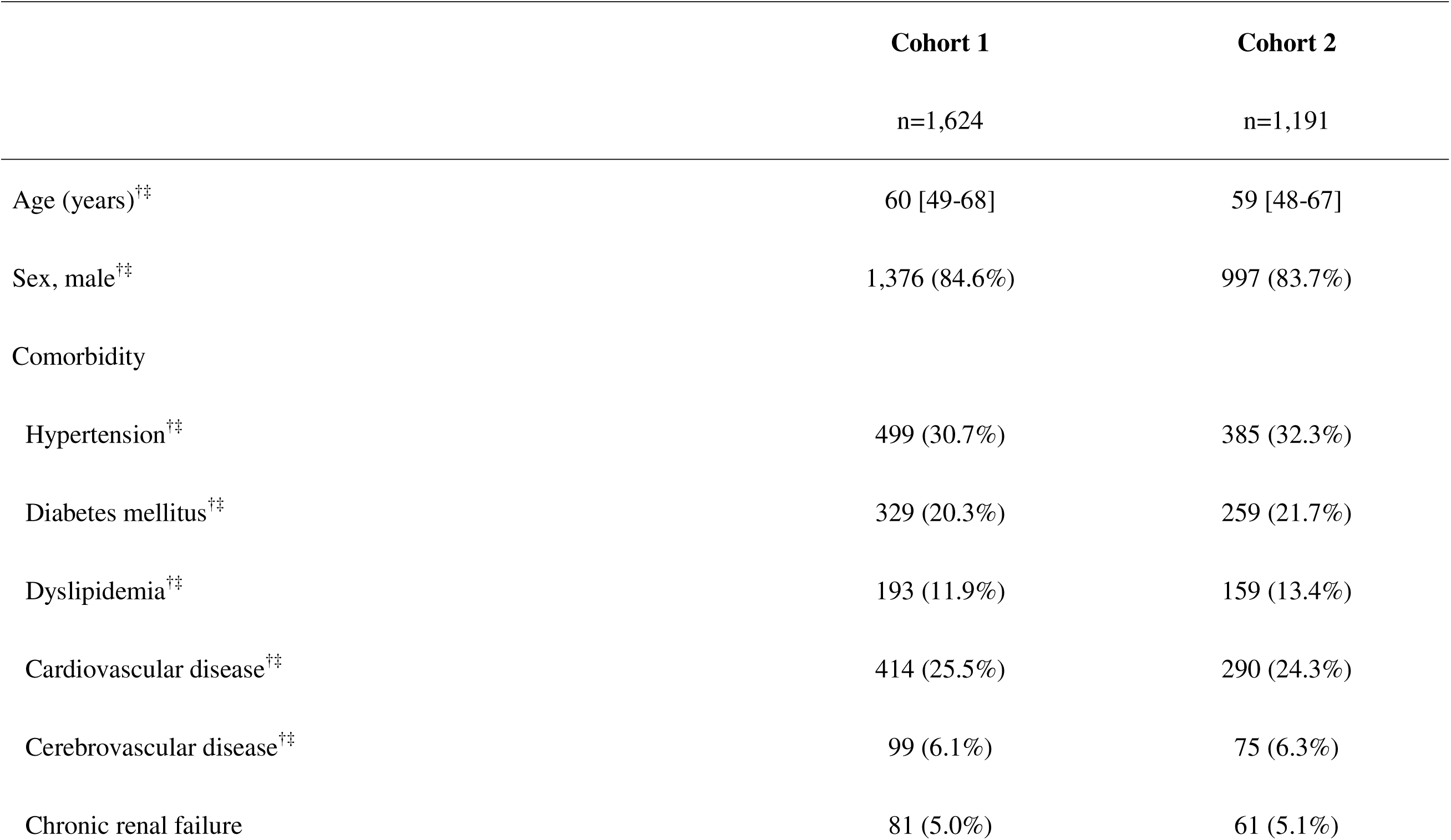
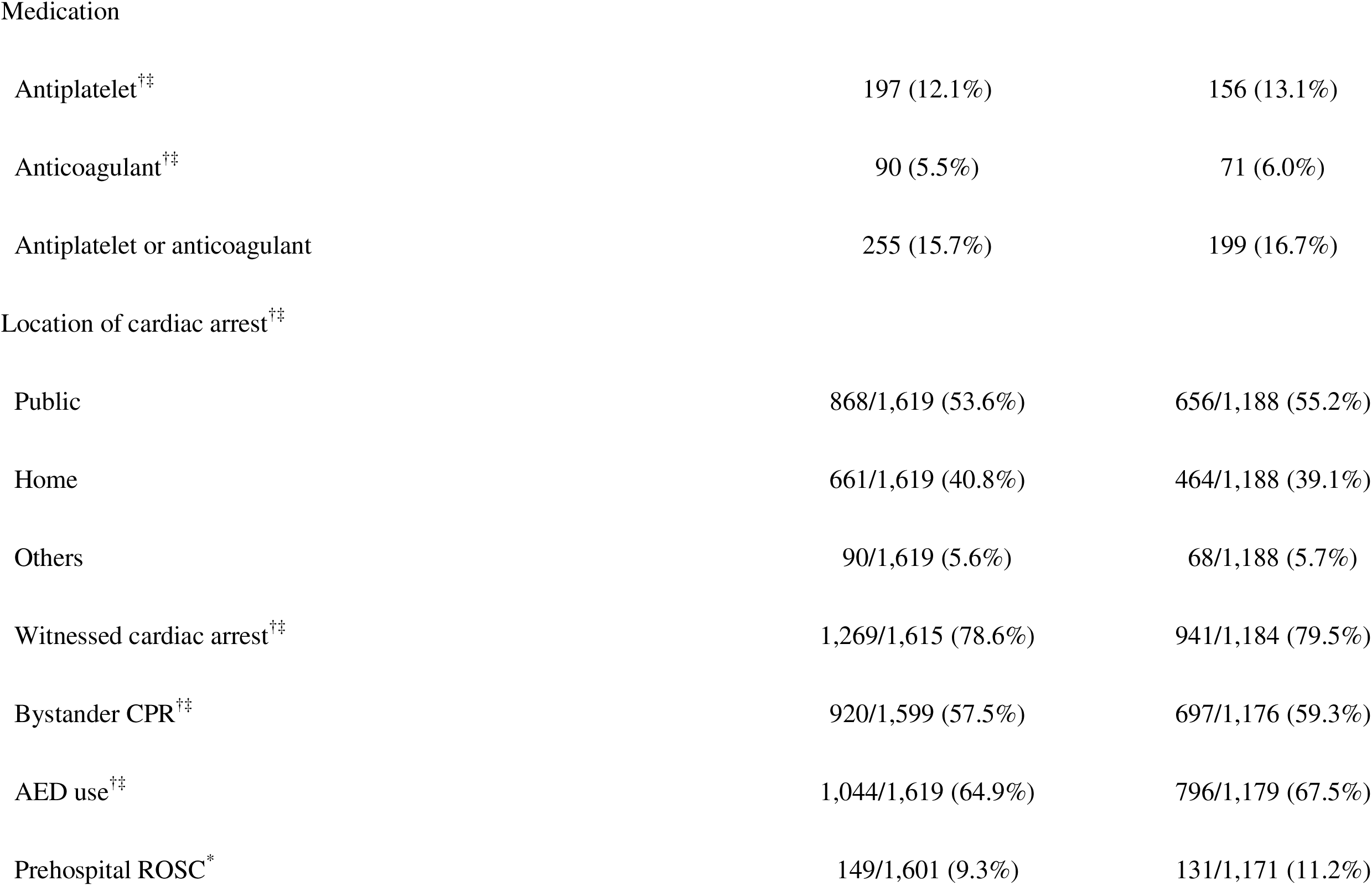
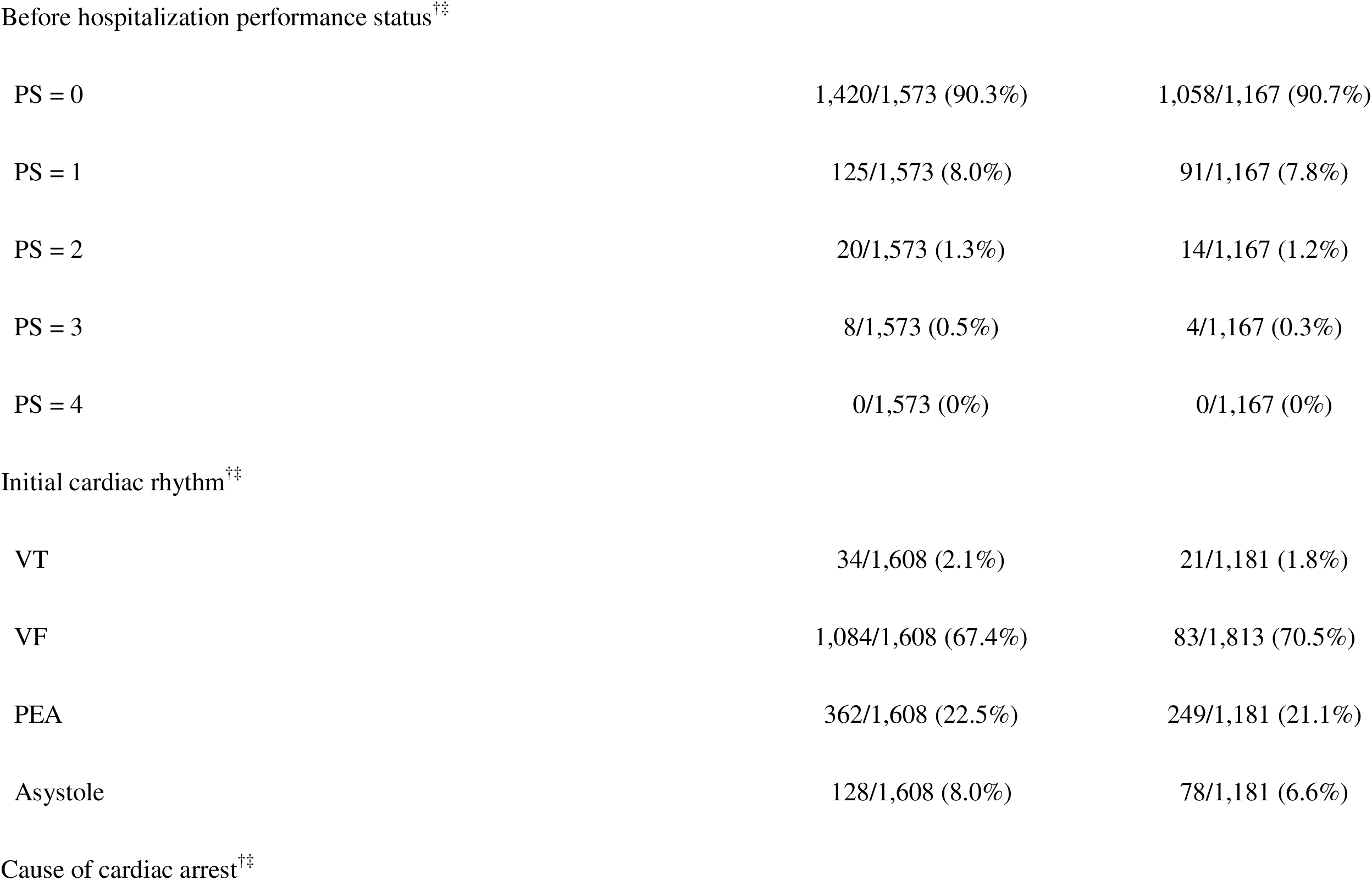
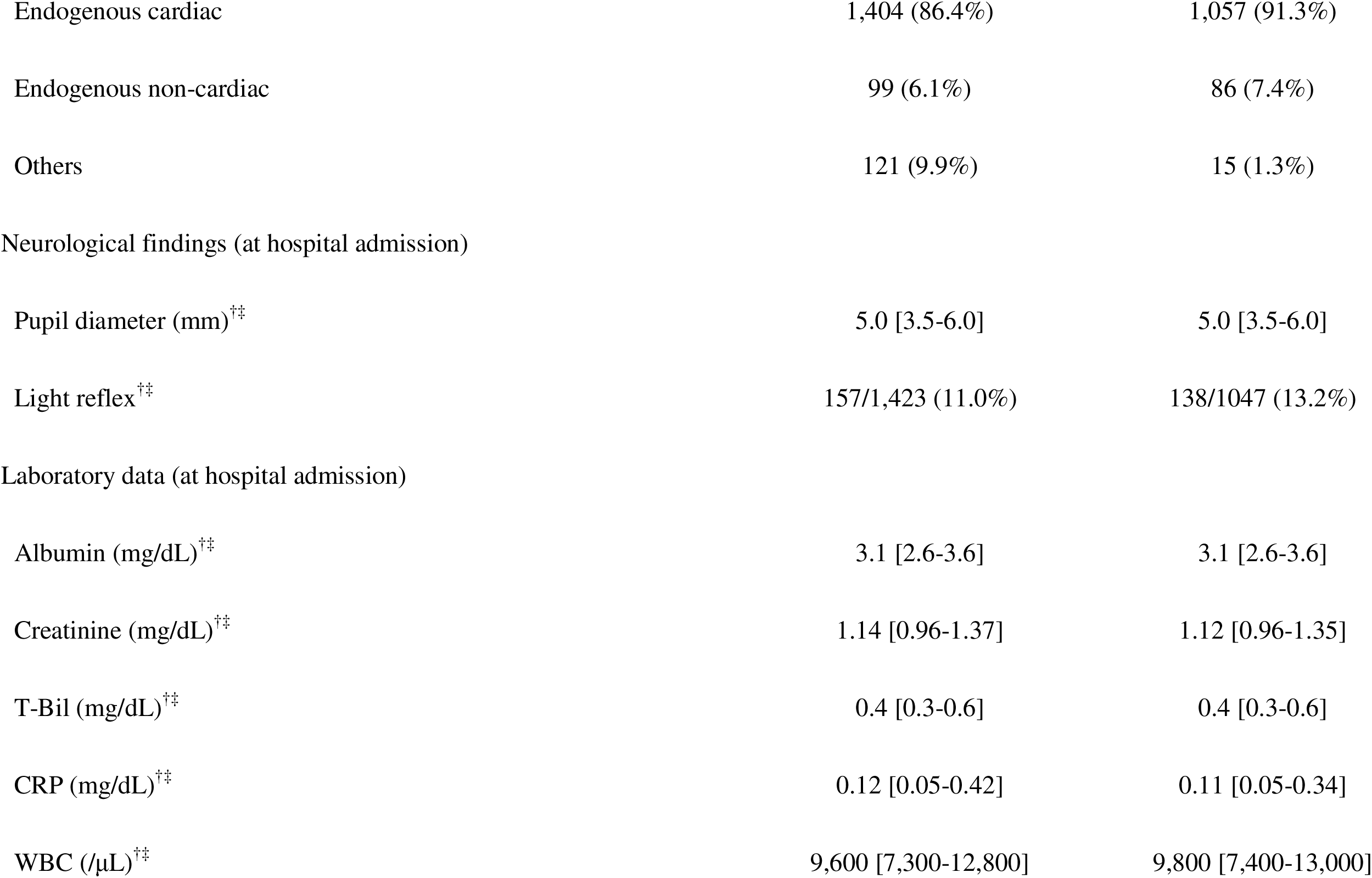
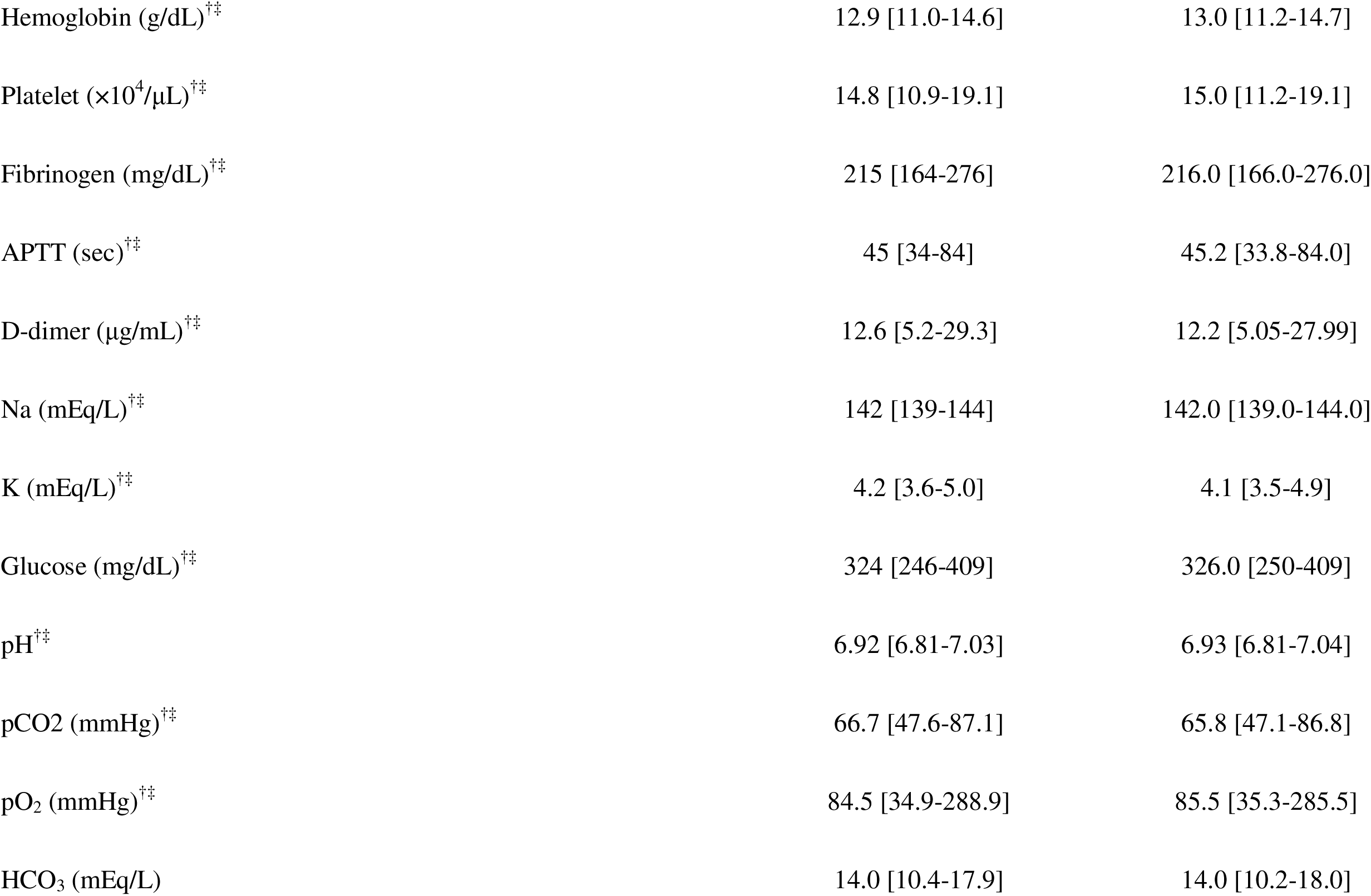
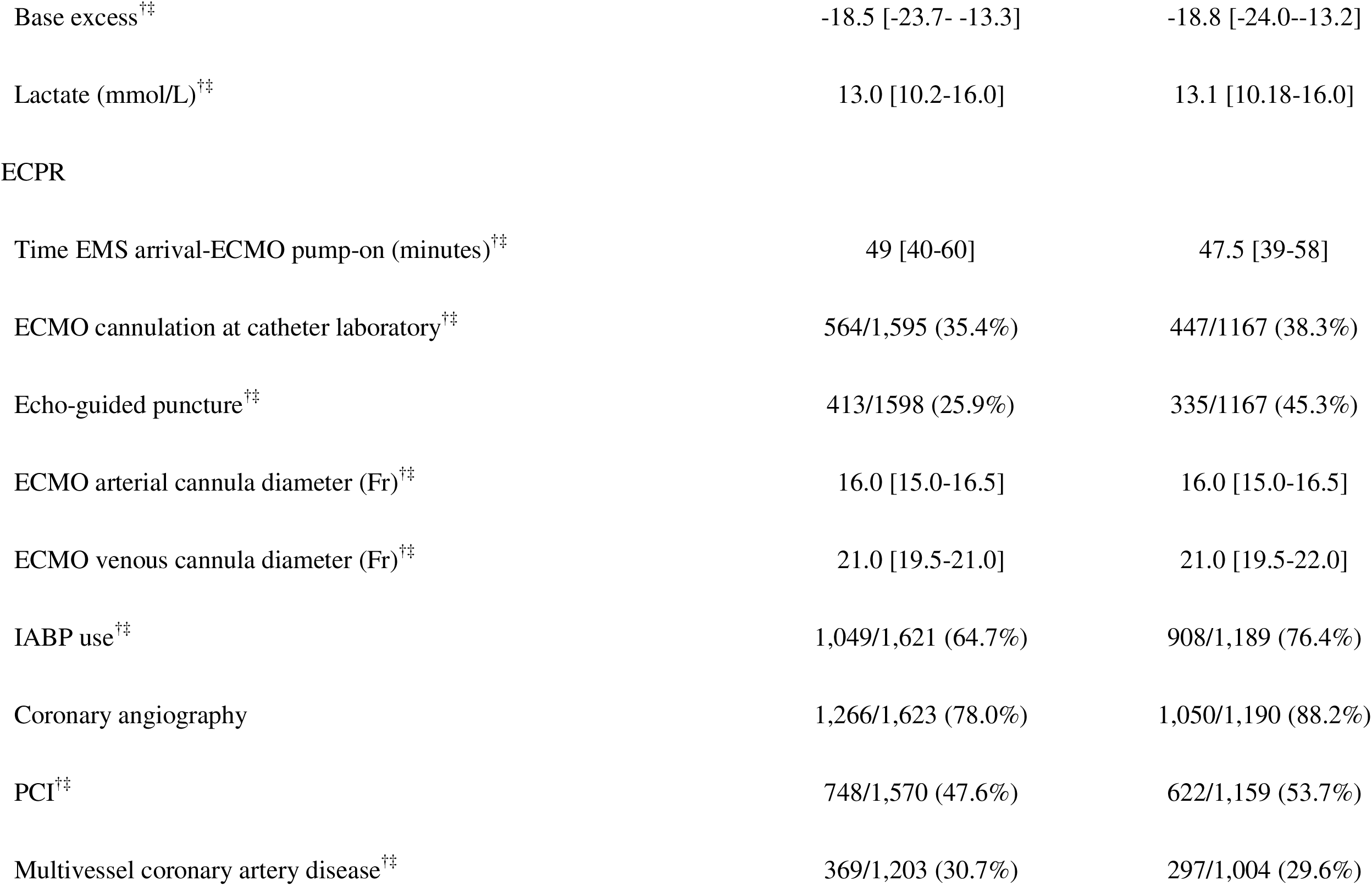
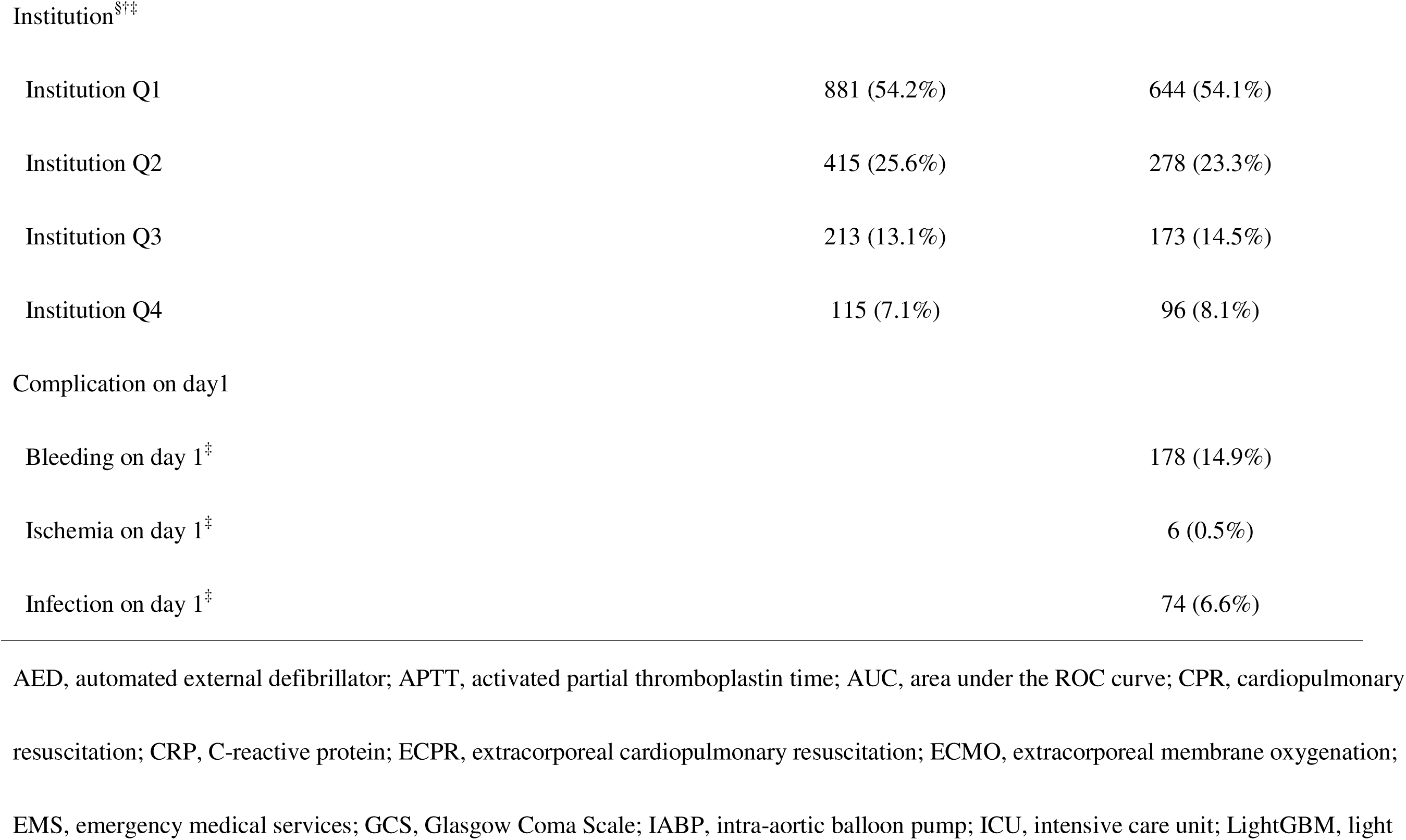
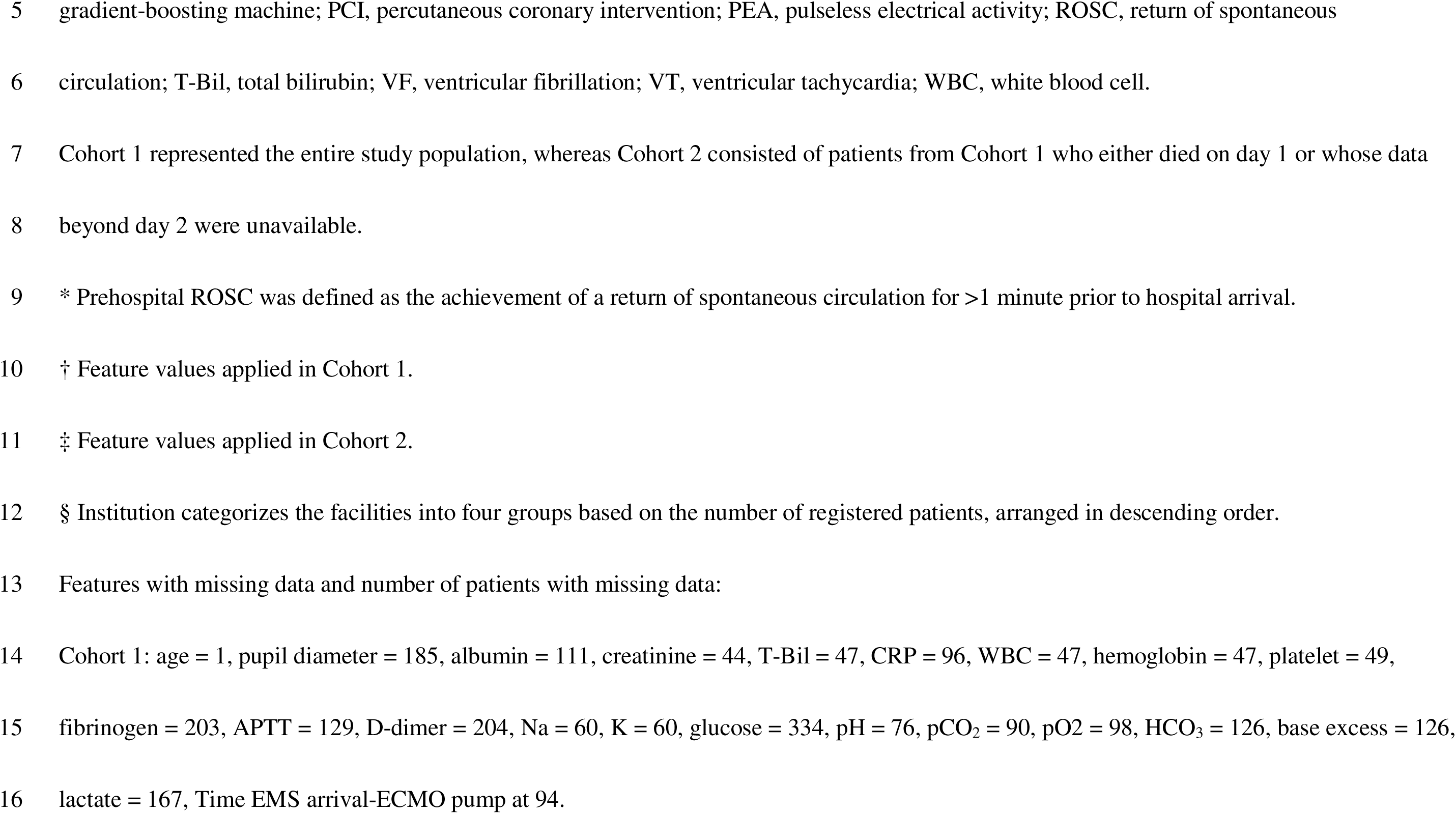
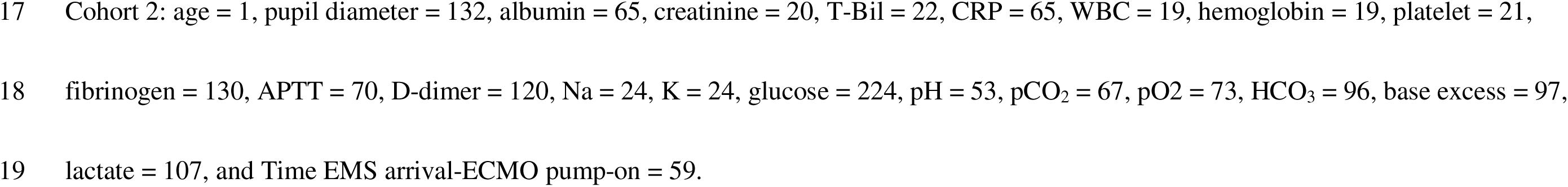
Baseline characteristics.

**Table 2:**
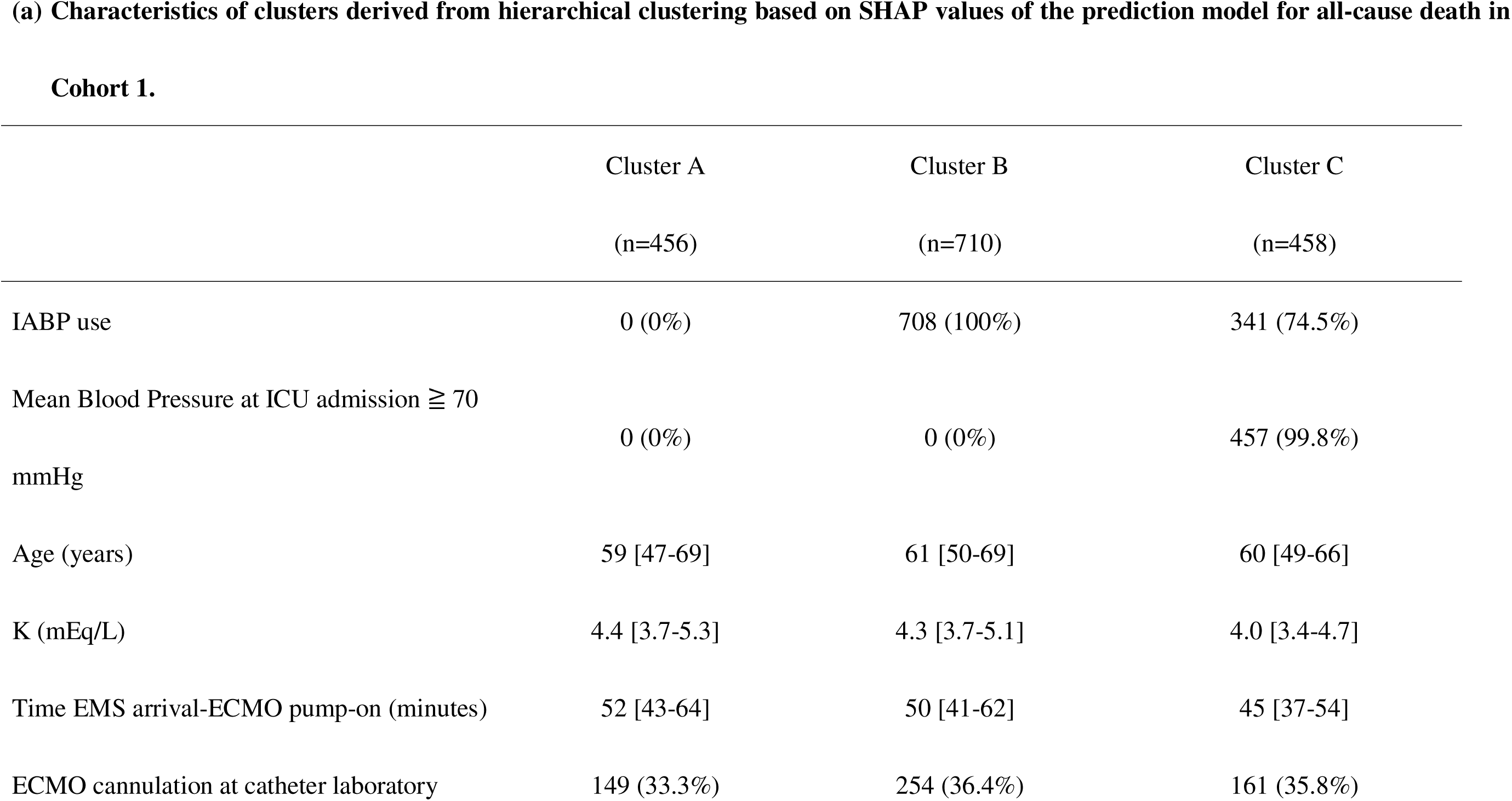
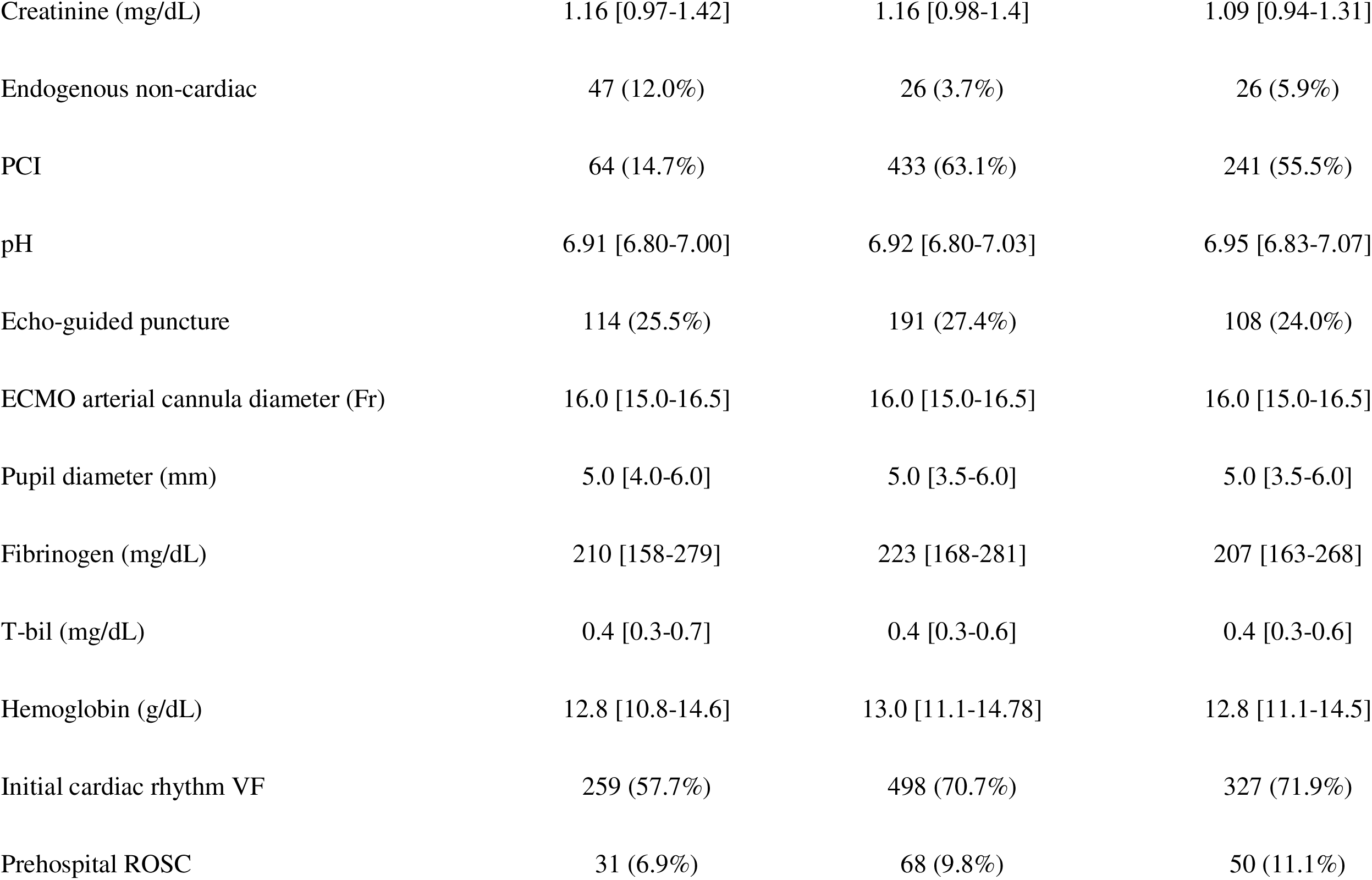
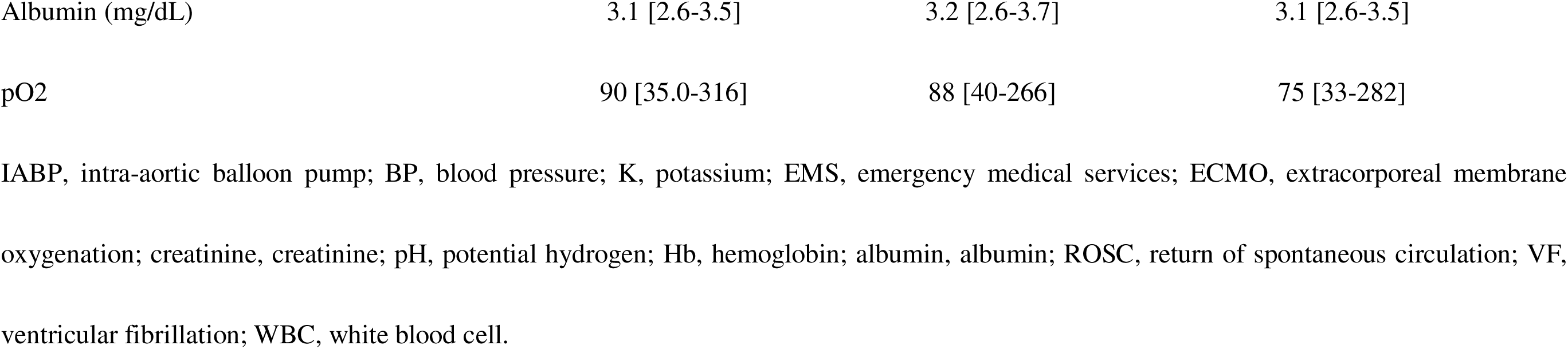
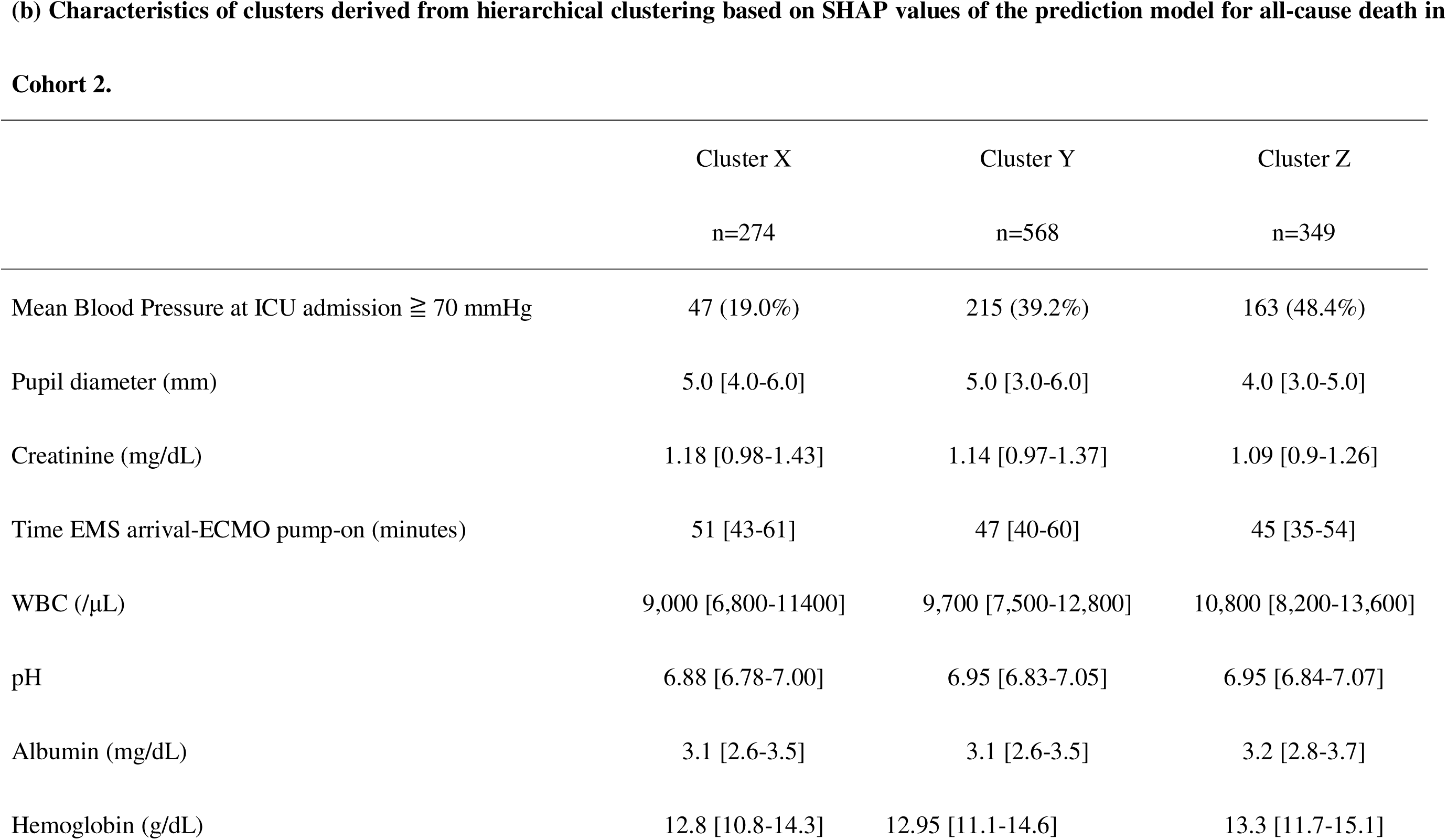
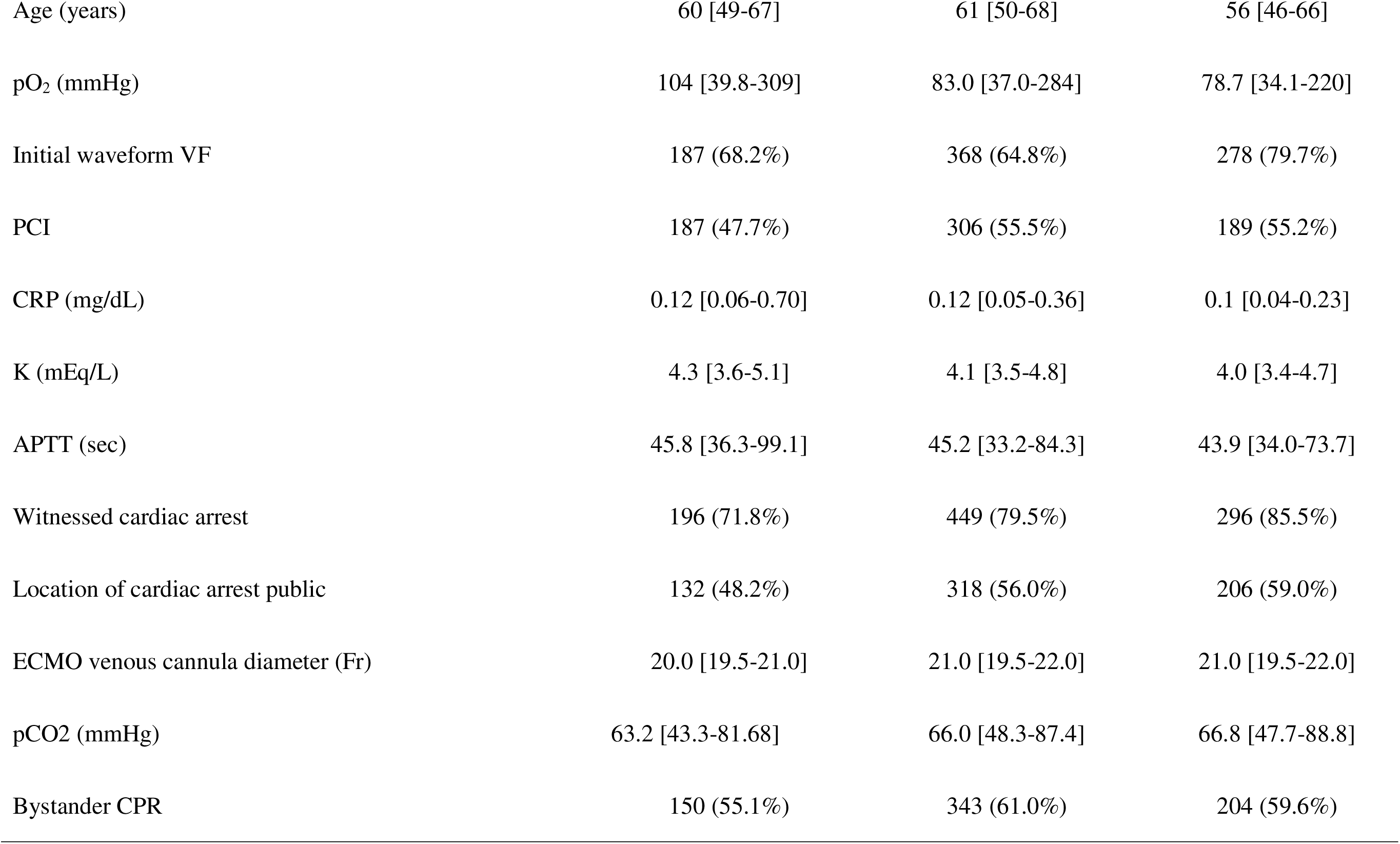
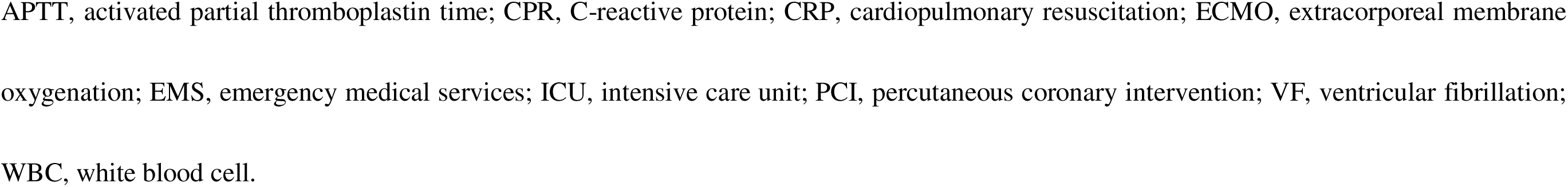
Characteristics of each cluster derived from SHAP-based clustering in each cohort.

## Discussion

In this multicenter cohort study of patients undergoing ECPR after OHCA, we developed XAI based prognostic models to predict early mortality and bleeding events. By applying a landmark analysis strategy at day 1, we successfully captured temporal differences in prognostic factors between the early phase and subsequent clinical course. The principal findings were: 1) the mortality prediction models demonstrated high discriminative performance across both timepoints, whereas the bleeding prediction models showed limited accuracy; 2) predictive variables contributing to mortality risk, as assessed by SHAP values, differed markedly between the early and later phases; 3) supervised clustering based on SHAP values effectively stratified patients into subgroups with distinct prognoses, which were not identifiable by traditional clustering based on raw clinical variables.

Prior studies have reported the utility of ML models in predicting outcomes after ECPR^8,9^. Kim et al. developed a ML model to predict neurological prognoses at discharge in 330 ECPR patients treated at a single institution, achieving an AUC of 0.84^8^. Similarly, Crespo-Diaz et al. constructed a ML model for patients with initial shockable rhythm, demonstrating an AUC of 0.80^9^. In comparison, our models achieved an AUC 0.85 for predicting day 1 mortality and a mean time-dependent AUC of 0.77 from day 2 onward. These performances are comparable or superior to those of previous studies. Moreover, unlike previous single-center studies, our work utilized a multicenter registry: SAVE-J II, thereby potentially offering greater generalizability and applicability to broader clinical settings.

A notable feature of our study is the identification of temporal heterogeneity in mortality risk factors. By implementing a landmark analysis at day 1, we developed two separate explainable AI models: one predicting events on day 1 in the overall cohort and another predicting subsequent events from day 2 among survivors. SHAP analysis revealed that key prognostic variables differed markedly between the two phases. Our SHAP-based interpretation revealed a temporal shift in key prognostic variables. On day 1, indicators of circulatory support and procedural timing, such as IABP use and the interval from EMS arrival to ECMO initiation, had high importance, whereas from day 2 onward, variables reflecting organ dysfunction, including creatinine and white blood cell count, became predominant. This observation suggests that the nature of mortality risk factors evolves during the early phase of post-ECPR care.

Notably, IABP use had the highest mean SHAP value in the day 1 model but was not among the top predictors thereafter. In Cohort 1, non-use of IABP was associated with higher predicted mortality. However, given that IABP use is subject to physician discretion, these findings must be interpreted cautiously. The SHAP values reflect the model’s prediction structure, not clinical causality. The preferential use of IABP in patients perceived to have a higher chance of survival may partially explain this association.

Supporting this, Naito et al. reported that decisions to withhold or withdraw life-sustaining therapy (WLST) were frequently made within the first two days after admission, suggesting that early deaths may be more influenced by clinical judgment than by irreversible pathology. In contrast, post-day 2 deaths may more accurately reflect the progression of organ failure.

The SHAP-based clustering approach further provided clinically meaningful patient stratification. By aggregating individualized feature attributions, we identified distinct subgroups with clearly different survival trajectories (Figures 5 and 6). In contrast, unsupervised clustering based on standardized raw clinical variables failed to achieve similarly clear prognostic separation (Supplemental Figures 1 and 2). SHAP values offer a unified scale across heterogeneous variables, emphasizing clinically informative features while minimizing the impact of irrelevant or redundant ones^21,22^. This attribute likely contributed to the superior performance of SHAP-based clustering compared to traditional methods. Prior studies have demonstrated the value of supervised clustering using SHAP in various settings, including acute heart failure, emergency department triage, and oncologic risk prediction^23–26^. Our findings suggest that SHAP-based clustering holds strong potential as a tool for risk-adapted management and prospective cohort stratification in the ECPR population.

**Figure 5: SHAP values in each cluster.**
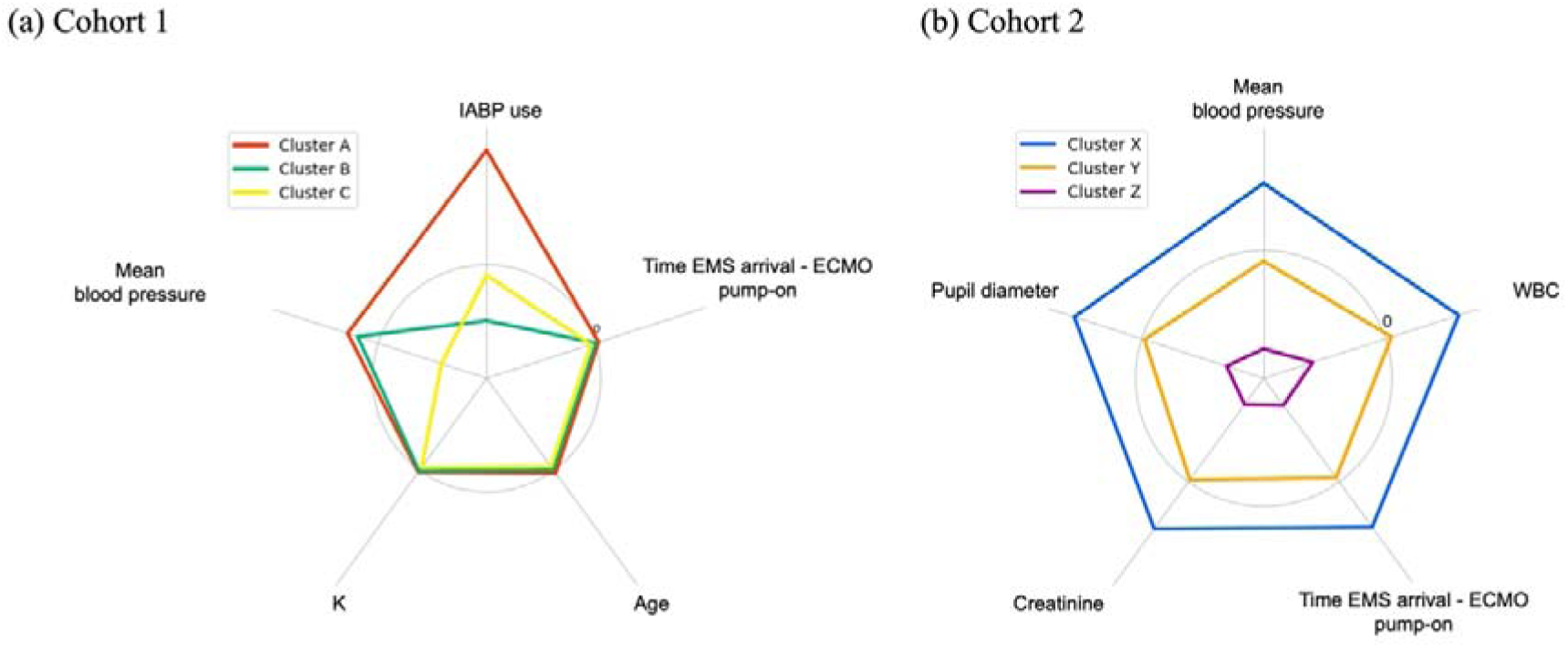
The mean SHAP values for each SHAP-based cluster are displayed in radar charts, with charts. (a) corresponding to Cohort 1 and (b) corresponding to Cohort 2. The SHAP value patterns indicated that in Cohort 1, the clusters were primarily differentiated by IABP use and blood pressure, whereas in Cohort 2, the clusters were distinguished by similarly correlated top-ranking variables.

**Figure 6: Cumulative density functions of clusters based on SHAP values for all-cause death.**
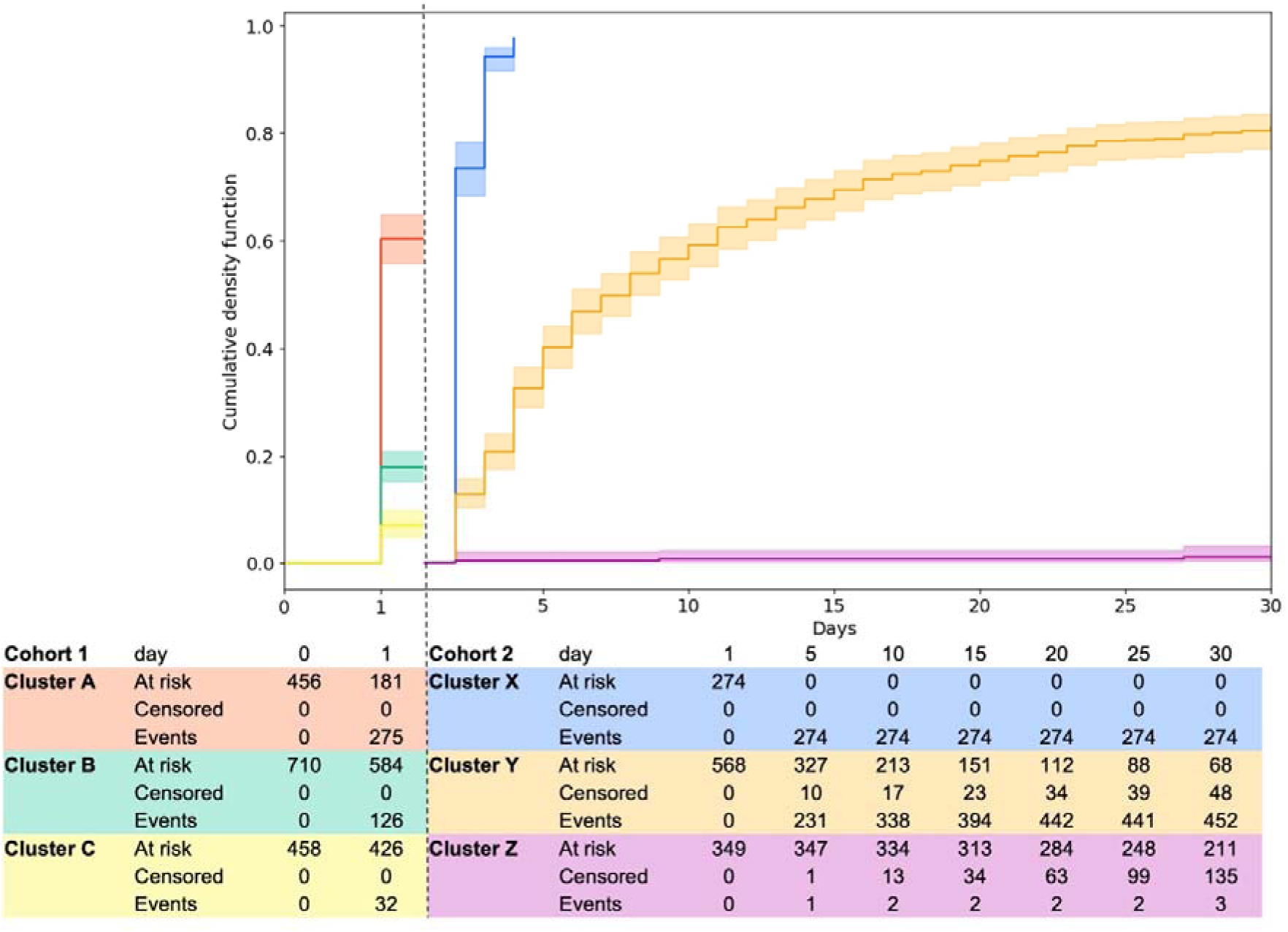
Cumulative density function at each time point for all-cause deaths for Cohort 1, clusters A, B, and C were defined by SHAP values from the LightGBM model predicting all-cause death on day 1. For Cohort 2, clusters X, Y, and Z were defined by the SHAP values from the gradient-boosting survival analysis model predicting all-cause death from day 2. The cumulative density functions for each cluster are also presented.

In contrast to the high performance in predicting all-cause mortality, our models demonstrated limited accuracy in predicting bleeding complications across both cohorts. This observation is consistent with prior reports in ECMO populations, where AI models have also successfully predicted mortality but struggled with complications such as acute brain injury^11,27^. Several factors may account for the limited predictive performance for bleeding events. First, bleeding encompasses diverse anatomical sites and varying severities, as noted by Isokawa et al.^28^, complicating uniform event characterization. Second, bleeding was broadly defined as events requiring transfusion or intervention, without central adjudication, introducing potential heterogeneity and misclassification bias. Additionally, institutional variation in the threshold for transfusion or intervention likely introduced additional noise. Together, these factors likely contributed to reduced model discriminative ability. Future work should consider stratifying bleeding events by anatomical site or severity grade to enhance predictive modeling.

This study had several limitations. First, it was a retrospective study, and external validation was not performed. Second, long-term prognostic data were unavailable. Third, there were no standardized treatment protocols, and the treatment strategies were determined individually. Fourth, in this registry, a significant proportion of cases underwent treatment withdrawal; however, all-cause deaths are treated equivalently for patients who died after withdrawal and those who died prior to withdrawal. ^1^ Fifth, the duration of cardiac arrest was unknown, and the EMS arrival to the ECMO pump-on time, which was used as a feature, was not equivalent to the low-flow time. Sixth, advances in treatment may have improved the outcomes, which may limit the applicability of the predictive model derived from the current registry. Finally, we did not evaluate other time points; however, there may be differences in inferential factors at other time points, as suggested in this study.

## Conclusions

In this multicenter study of OHCA patients undergoing ECPR, we developed AI models to predict early mortality and bleeding complications. Our models demonstrated high discriminative performance for mortality prediction, while prediction of bleeding events remained challenges. SHAP-based interpretation revealed distinct temporal shifts in key prognostic factors between the early and later phases of care. Furthermore, SHAP-based supervised clustering successfully stratified patients into subgroups with clinically meaningful differences in outcomes. These findings highlight the potential of explainable AI not only for accurate prediction but also for interpretable risk stratification, supporting personalized clinical decision-making in the management of ECPR-treated patients.

## Acknowledgments

We acknowledge Editage (www.editage.jp) for their assistance with English language editing. We would like to thank all the members of the SAVE-J II study group who participated in this study: Hirotaka Sawano, M.D., Ph.D. (Osaka Saiseikai Senri Hospital), Yuko Egawa, M.D., Shunichi Kato, M.D. (Saitama Red Cross Hospital), Kazuhiro Sugiyama, M.D., Maki Tanabe, M.D. (Tokyo Metropolitan Bokutoh Hospital), Naofumi Bunya, M.D., Takehiko Kasai, M.D. (Sapporo Medical University), Shinichi Ijuin, M.D., Shinichi Nakayama, M.D., Ph.D. (Hyogo Emergency Medical Center), Jun Kanda, M.D., Ph. D., Seiya Kanou, M.D. (Teikyo University Hospital), Toru Takiguchi, M.D., Shoji Yokobori, M.D., Ph.D. (Nippon Medical School), Hiroaki Takada, M.D., Kazushige Inoue, M.D. (National Hospital Organization Disaster Medical Center), Ichiro Takeuchi, M.D., Ph.D., Hiroshi Honzawa, M.D. (Yokohama City University Medical Center), Makoto Kobayashi, M.D., Ph.D., Tomohiro Hamagami, M.D. (Toyooka Public Hospital), Wataru Takayama, M.D., Yasuhiro Otomo, M.D., Ph.D. (Tokyo Medical and Dental University Hospital of Medicine), Kunihiko Maekawa, M.D. (Hokkaido University Hospital), Takafumi Shimizu, M.D., Satoshi Nara, M.D. (Teine Keijinkai Hospital), Michitaka Nasu, M.D., Kuniko Takahashi, M.D. (Urasoe General Hospital), Yoshihiro Hagiwara, M.D., M.P.H. (Imperial Foundation Saiseikai, Utsunomiya Hospital), Shigeki Kushimoto, M.D., Ph.D. (Tohoku University Graduate School of Medicine), Reo Fukuda, M. D. (Nippon Medical School Tama Nagayama Hospital), Takayuki Ogura, M.D., Ph.D. (Japan Red Cross Maebashi Hospital), Shin-ichiro Shiraishi, M.D. (Aizu Central Hospital), Ryosuke Zushi, M.D. (Osaka Mishima Emergency Critical Care Center), Norio Otani, M.D. (St. Luke’s International Hospital), Migaku Kikuchi, M.D., Ph.D. (Dokkyo Medical University), Kazuhiro Watanabe, M.D. (Nihon University Hospital), Takuo Nakagami, M.D. (Omihachiman Community Medical Center), Tomohisa Shoko, M.D., Ph.D. (Tokyo Women’s Medical University Medical Center East), Nobuya Kitamura, M.D., Ph.D. (Kimitsu Chuo Hospital), Takayuki Otani, M.D. (Hiroshima City Hiroshima Citizens Hospital), Makoto Aoki, M.D., Ph.D. (Gunma University Graduate School of Medicine), Masaaki Sakuraya, M.D., M.P.H. (JA Hiroshima General Hospital Hiroshima), Hideki Arimoto, M.D. (Osaka City General Hospital), Koichiro Homma, M.D., Ph.D. (Keio University School of Medicine), Hiromichi Naito, M.D., Ph.D. (Okayama University Hospital), Shunichiro Nakao, M.D., Ph.D. (Osaka University Graduate School of Medicine), Tomoya Okazaki, M.D., Ph.D. (Kagawa University Hospital), Yoshio Tahara, M.D., Ph.D. (National Cerebral and Cardiovascular Center), Hiroshi Okamoto, M.D, M.P.H. (St. Luke’s International Hospital), Jun Kunikata, M.D., Ph.D., Hideto Yokoi, M.D., Ph.D. (Kagawa University Hospital).

## Data availability statement

The data underlying this article are subject to restrictions and cannot be shared publicly.

## Non-standard Abbreviations and Acronyms

AI: Artificial intelligence
ECPR: Extracorporeal cardiopulmonary resuscitation
ELSO: Extracorporeal Life Support Organization
EMS: Emergency medical service
EST: Extra survival trees
GBSA: Gradient-boosting survival analysis
IABP: Intra-aortic balloon pumping
IQR: Interquartile range
LightGBM: Light Gradient-Boosting machine
OHCA: Out-of-hospital cardiac arrest
PCI: Percutaneous coronary intervention
RF: Random forest
ROC: Receiver operating characteristic
ROSC: Return of spontaneous circulation
RSF: Random survival forest
SHAP: Shapley Additive Explanations
WBC: White blood cell

