## supplemental materials for "Explainable Artificial Intelligence for Prognostic Stratification in Out-of-Hospital Cardiac Arrest Patients Undergoing Extracorporeal Cardiopulmonary Resuscitation"

**Supplemental Figure 1**
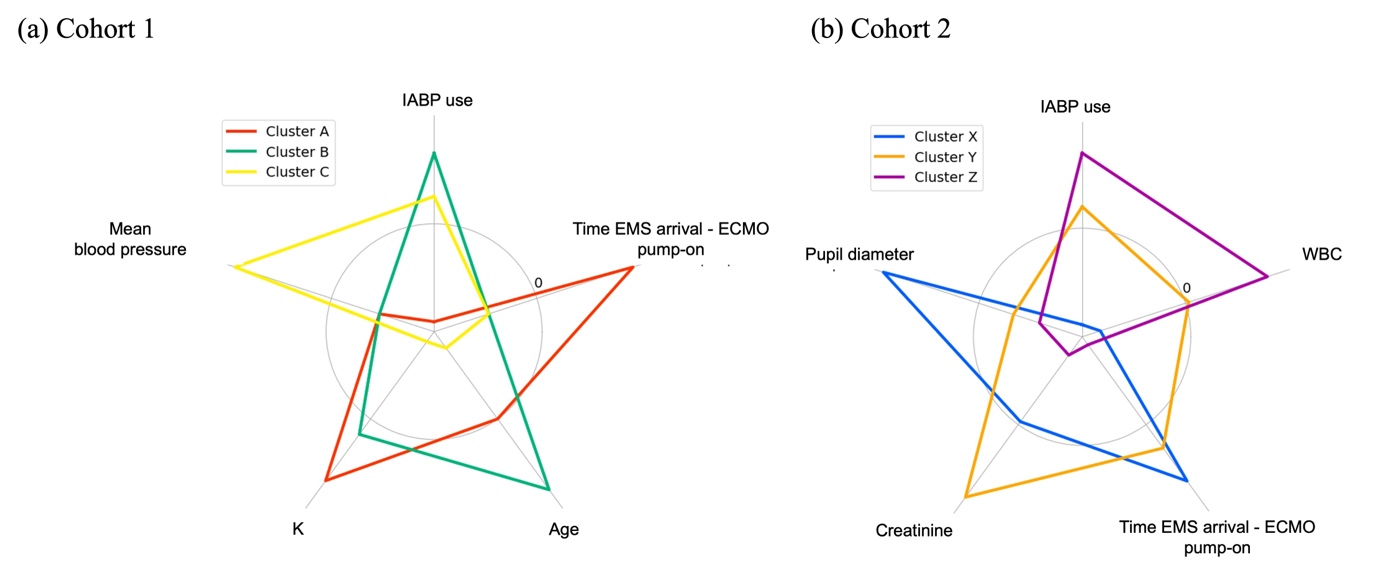

**Supplemental Figure 2**
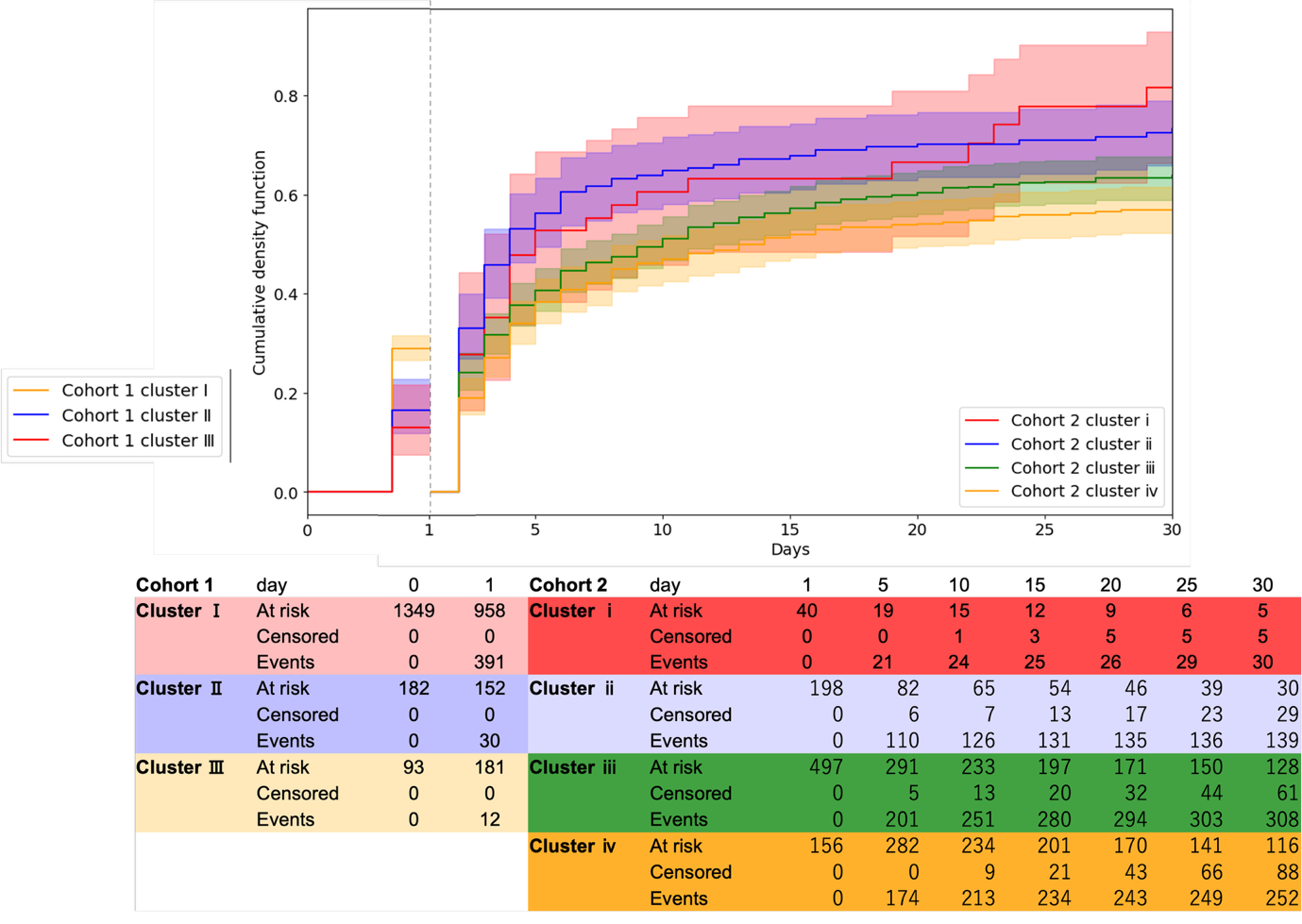

**Supplemental Figure legends**

**Supplemental Figure 1: Standardized observed values or proportions for each cluster derived from SHAP-based clustering.** Radar charts depict the standardized or proportion of observed values for each cluster derived from SHAP-based clustering, with chart (a) corresponding to Cohort 1 and chart (b) corresponding to Cohort 2.

**Supplemental Figure 2: Cumulative density functions of clusters based on standardized observed values for all-cause death.** Cumulative density function at each time point for all-cause mortality For Cohort 1, clusters I, II, and III were defined using hierarchical clustering based on standardized values. In Cohort 2, clusters i, ii, iii, and iv were defined using the same method. The cumulative density functions for each cluster are also presented.
